# Super-spreaders of novel coronaviruses that cause SARS, MERS and COVID-19 : A systematic review

**DOI:** 10.1101/2022.03.14.22272351

**Authors:** Julii Brainard, Natalia R. Jones, Florence Harrison, Charlotte C. Hammer, Iain R. Lake

## Abstract

**OBJECTIVE:** Most index cases with novel coronavirus infections transmit disease to just 1 or 2 other individuals, but some individuals ‘super-spread’ – they are infection sources for many secondary cases. Understanding common factors that super-spreaders may share could inform outbreak models.

**METHODS:** We conducted a comprehensive search in MEDLINE, Scopus and preprint servers to identify studies about persons who were each documented as transmitting SARS, MERS or COVID-19 to at least nine other persons. We extracted data from and applied quality assessment to eligible published scientific articles about super-spreaders to describe them demographically: by age, sex, location, occupation, activities, symptom severity, any underlying conditions and disease outcome. We included scientific reports published by mid June 2021.

**RESULTS:** The completeness of data reporting was often limited, which meant we could not identify traits such as patient age, sex, occupation, etc. Where demographic information was available, for these coronavirus diseases, the most typical super-spreader was a male age 40+. Most SARS or MERS super-spreaders were very symptomatic and died in hospital settings. In contrast, COVID-19 super-spreaders often had a very mild disease course and most COVID-19 super-spreading happened in community settings.

**CONCLUSION:** Although SARS and MERS super-spreaders were often symptomatic, middle- or older-age adults who had a high mortality rate, COVID-19 super-spreaders often had a mild disease course and were documented to be any adult age (from 18 to 91 years old). More outbreak reports should be published with anonymised but useful demographic information to improve understanding of super-spreading, super-spreaders, and the settings that super-spreading happens in.

## Introduction

During the 2020 Covid-19 pandemic the role of super-spreaders in the transmission of the disease became a widely discussed topic in the media, and within the scientific community. Covid super-spreading was publicised in newspapers during 2020 from fitness classes, religious worship, skiing trips and public transport. Infectious disease super-spreader has long been recognised. A long-standing rule of thumb is that 20% of the host population give rise to 80% of the transmission potential (20/80 rule) (Woolhouse *et al*. 1997). Super-spreading was initially best described across a range of vector-borne and sexually transmitted diseases (ibid), while more recent research described frequent super-spreading among the novel coronaviruses (nCoV) that cause SARS (Stein 2011), MERS (Kucharski & Althaus 2015) and COVID-19 (Frieden & Lee 2020). Fully understanding the role of super-spreaders could enable more effective containment of disease outbreaks, and more accurate modelling of epidemics. There have been descriptions of super-spreading events for SARS (Shen *et al*. 2004) and MERS (Kang *et al*. 2017), while documentation of super-spreading from the COVID-19 pandemic is still emerging. However, to date there is very little focus on the characteristics of the individual super-spreader. By compiling details of super-spreading events across the three novel coronavirus outbreaks, we aimed to assess whether there are any common factors between the super-spreaders themselves that may aid early identification, perhaps prevent super-spreading, and therefore help to reduce total transmission. Given the 20/80 rule, identifying, containing and treating potential super-spreaders early may help reduce harms from future pandemics (Stein 2020, Kain *et al*. 2021).

Although super-spreaders became high profile cases in the media (e.g. ‘Patient 31’ in South Korea (Kasulis 2020) (Reuters 2020) and the Alps cluster (Danis *et al*. 2020b)) they usually represent a very small proportion of the total number of disease cases. During the Singapore SARS outbreak of 2003 it was noted that among the first wave of cases, the vast majority (81%) had not passed the disease on to any other cases (Leo *et al*. 2003), while 5 people (2.5%) had spread the infection to 10 or more people each (Lipsitch *et al*. 2003).

Reasons offered to explain why only some cases become super-spreaders can be grouped into four categories: pathogen (the causes of the illness and the means by which it can be transmitted) biological/individual (e.g. age, sex, ethnicity, viral load, shedding), behavioural (e.g. type of work, number of people they come into contact with), and environmental (e.g. air circulation in work place) (Frieden & Lee 2020). However, the relative importance may vary by disease type e.g. sexually transmitted disease vs vector-borne vs respiratory, and possibly between individual pathogens (e.g. between different coronaviruses).

Focusing on individual profiles is complimentary to identifying contexts when super-spreading happens. Two questions posed by Stein (2011; pg e512) still need to be answered for COVID-19 specifically and coronaviruses generally:

> “What makes certain hosts become super-spreaders, and how can we identify them in a timely manner?”

Identifying super-spreading individuals or events is challenging, as without detailed contact tracing or genetic testing of the virus it is difficult to make definite links between index and secondary cases. Links typically must be made by assessing potential exposure windows and elapsed time until the start of symptoms. Many asymptomatic cases may be missed. A better understanding of the characteristics of super-spreaders may help identify super-spreader events, and therefore further assist in our ability to prevent them. Better public understanding of the risks that possible super-spreaders pose may also help others to recognise their own exposure or potential contribution to super-spreading, and thus improve compliance with public health guidelines.

To this end, we undertook this review to research attributes of individuals believed to be index patients in super-spreading events of novel coronaviruses (MERC, SARS, COVID-19) during June 2021. We were interested in factors like sex, age, severity of symptoms, disease course outcomes, transmission setting, ethnicity, occupation and distribution of count of cases in cluster.

## Methods

We defined a “super-spreader” as an index case who was described in the scientific literature as having caused at least nine secondary infections. We chose this threshold (nine) to be indisputably much higher than the commonly cited effective reproductive numbers (around 3.0) for all of the novel coronavirus diseases: SARS, MERS, and (initially) COVID-19 (World Health Organization 2003, Choi *et al*. 2018, Viceconte & Petrosillo 2020)). There is no formal consensus on how to define super-spreaders relative to typical reproductive number for a stated pathogen (Al-Tawfiq & Rodriguez-Morales 2020). The WHO defined super-spreading as “above the average number of secondary cases” for SARS (World Health Organization 2003). Others suggested that super-spreaders should be defined as the 1% of index cases that generated the most secondary infections (Lloyd-Smith *et al*. 2005, Xu *et al*. 2020b). We acknowledge that these other definitions of ‘super-spreader’ have merits.

We were only interested in transmission between humans outside of deliberate experiments. We refer to ‘super-spreading events’ as events where super-spreading is believed to have happened. Events could conceivably range from (exemplars) a small birthday party to the annual Hajj in size and scope.

There was ambiguity in reports about how many secondary infections were directly caused by each index case. Different reports identified different numbers of secondary cases, sometimes fewer than nine. We tried to be inclusive about this eligibility criteria: if at least one peer-reviewed assessment identified at least nine secondary infections, we included the individual as a super-spreader. We report below the full range of possible counts of secondary cases from each specific index case as identified in relevant literature even if some of these were less than nine cases.

We undertook a structured search of scientific bibliographic databases using the search terms in Box 1. The initial searches were run on 3 June 2020 with an updated search on 18 June 2021. The search results were combined into a single database and de-duplicated. This review has PROSPERO registration CRD42020190596.

### Screening

Eligible publications were published in 2002 or later. Our latest search date was 18 June 2021. We considered publications in all languages if we could translate them to English or Spanish. Two independent screeners applied eligibility criteria, with a third person to decide if no consensus. Articles had to be chosen by at least two reviewers to go to full text review. Full-text review confirmed eligibility criteria. The report had to describe infection(s) confirmed by rt-PCR, cell-culture or clinical presentation and known exposure history. Eligibility criteria were:

- Report must describe one of these 3 novel coronavirus (nCoV) diseases that primarily spread by respiratory face to face transmission: MERS, SARS, COVID-19
- Study design: almost any, but case or case-cluster reports were preferred. Preprints or grey literature from credible sources could be used and could be supported by sources such as public statements by individuals themselves, interviews and press releases.
- Any unplanned non-laboratory setting (e.g., clinical, schools, homes, religious, etc.).

Box 1. Phrases used for scientific bibliographic database searches

**SCOPUS** SEARCH

(TITLE-ABS-KEY (coronavirus OR covid* OR wuhan OR mers* OR “Middle East Respiratory” OR sars OR “sudden acute

respiratory”) AND ALL (super*)) AND PUBYEAR > 1999 AND (LIMIT-TO (SUBJAREA, “MEDI”))

**medrxiv/bioRxiv/Preprint.org** servers

(coronavirus or COVID* or MERS* or “Middle East Respiratory” or SARS* or “sudden acute respiratory syndrome”)

And

Super*

**OVID MEDLINE & EMBASE**

The search phrases was

((coronavirus or COVID* or MERS* or “Middle East Respiratory” or SARS* or “sudden acute respiratory syndrome”).ti. or

((coronavirus or COVID* or MERS* or “Middle East Respiratory” or SARS* or “sudden acute respiratory syndrome”).kw. or

((coronavirus or COVID* or MERS* or “Middle East Respiratory” or SARS* or “sudden acute respiratory syndrome”).ab.)

and

((super*).tx.)

### Extraction and Synthesis

We used spreadsheets to record specific extracted details about the index case, the setting of their super-spreading activities and their contacts. One investigator undertook initial extractions which were checked by another researcher, with differences resolved by discussion or a third opinion. We then tabulated and descriptively summarised this information, separately for each disease. The extracted information was grouped by disease (MERS, SARS, COVID-19). Pooled summary was only undertaken if there were at least 20 super-spreaders found for any disease. No pooling of data beyond descriptive statistics (such as median age or percentage female) was attempted for smaller groups.

### Activities when super-spreading

We knew that most MERS and SARS super-spreaders were clinical inpatients and their occupation did not influence their super-spreading. We reported in summary format the activities that index spreaders seem to have been doing (such as work in a call centre or attendance to a dance fitness class, etc). The information was expected to be heterogeneous and reported as stated by authors.

### Age and sex of the super-spreader

This was extracted as recorded in years, and reported by disease. The information was listed in tabular format (one row per super-spreader) if there were less than 20 super-spreaders associated with a disease. When there were at least 20 super-spreading individuals (per disease) information was reported in graphical format, and included medians, interquartile range and 5% & 95% confidence intervals, further broken down by MERS/SARS/COVID-19. If there were at least 20 eligible index patients, we discussed whether the age distribution of super-spreaders was representative of the wider community population.

### Ethnicity

We knew from preliminary searches that this information is largely unavailable, but collected the information we found and reported it narratively. The information was heterogeneous and diverse and sometimes non-specific such as ‘Chinese’ or ‘African American’.

### Occupation

We knew from preliminary searches that this information is largely unavailable, but narratively reported the information we found. The information was expected to be heterogeneous and was reported as stated by authors.

### Secondary cases

We recorded the direct count of first generation cases and took this to equal the number of persons believed that the super-spreader infected. This was expressed as individual case counts. This estimate was sometimes described as a range and was often reported as different numbers in different articles. We reported available information (including ranges of estimates) as stated by original authors.

### Setting

We determined where the super-spreader had their impact, in settings such as households, hospitals or churches, etc.. Additionally, the country was reported, as was the city or other regional information when available. The information was heterogeneous and reported as stated by authors.

We intended to group settings where this was potentially informative, into sets such as ‘workplaces’, ‘schools’, ‘military ships’ or ‘pleasure cruises’. For reporting purposes setting may be grouped with activities when super-spreading.

### Symptoms and disease outcomes

Outcomes (especially mortality rates) for the super-spreaders were summarised in simple narrative format.

### Underlying conditions

We recorded narratively which underlying conditions the super-spreader individuals were reported to have (such as diabetes or heart disease). There is a hypothesis that super-spreaders are more likely (than the general population) to be persons with compromised immune systems (Lakdawala & Menachery 2021). Conversely, there was also widespread public perception early in the COVID-19 pandemic (Grey 2020) that super-spreaders might often be nearly or completely asymptomatic. We collected data that might address either hypothesis.

### Deviations from protocol

We had planned to report on Tertiary and onward transmission counts, broken into percentiles or median estimates. We extracted this information but found that it was reported with such great variability and inconsistency that it was not possible to summarise, and is not elaborated upon further.

### Quality Assessment

We designed a customised quality assessment of the reporting sources, see Table 1. This checklist was designed to assess the credibility of the sources and the rigour of their contact tracing methods and transmission chain reconstructions. We defined the “Most credible” studies to be those with Yes answers to all six questions, studies with 4-5 Yes answers as “Fairly credible” and those with three or fewer Yes answers as “Less credible”.

**Table 1.** Quality Assessment Checklist for included studies

|  | Category | Answers (from cluster of sources about each event) |
| --- | --- | --- |
| 1.Primary | Is the document a primary source of information, i.e. have they conducted the study themselves or are they reporting on the work of others? | YES: collected majority of info themselves, but may mention some secondary data to inform their summary. Primary needs to be original interviews or review of original medical records.<br>NO: Obviously mostly or entirely citing others<br>UNCLEAR: other answers, including unclear sources or not clear that majority of the info being reported was primary data collection |
| 2.Document | Was main source of information been subject to peer review? | YES: Publication title = in Scimago or Pubmed<br>NO: other source |
| 3.Case definition | Is there a formal case definition (ideally chain based upon a PCR/antigen/lab test among at least one of the index/secondaries) as opposed to only symptoms etc. *If not explicitly stated this could be surmised if the report from a health agency.* | YES: There is a statement what case definition is, and this involves either<br>A) Contact history/chain with lab-confirmed cases and fever+cough symptoms (don't strictly define fever)<br>B) PCR- or antigen-confirmation of most cases<br>NO: No case definition provided<br>UNCLEAR: Other case definition |
| 4.Rigour | Do sources state that a formal contact tracing procedure was in place? *If not explicitly stated this could be surmised if the report is from a health agency.* | YES: Describes interview(s) with most/all original index case(s)<br>NO: clearly No Interviews with index cases or valid proxies<br>UNCLEAR: Few/minority interviews with index/contacts, or can't tell, or not easy to answer YES or NO |
| 5.Exposure | Is there evidence that there has been close (enough) contact between the super-spreader and infected individuals (e.g. same hospital ward, apartment) **This may be the trickiest to define** | YES: Is plausible from contact history that had opportunity to spend at least 10 minutes within 2m of each other; or another mechanism for super-spreading has been suitably described and defended (eg., Amoy Gardens aerosolised)<br>NO: Contact history not describing plausible high risk contact opportunit(ies)<br>UNCLEAR: If contact history not sufficiently described |
| 6.Timeliness | Do the dates of contacts and illness match likely incubation periods (i.e. MERS median 5 range 2-14 days, SARS variable 2-7 days, COVID median 5 range 2-14 days) | YES: see windows <-<br>NO: outside those windows<br>UNCLEAR: not stated, can't be sure |
| Scoring: number of YES answers: Most credible: 6 YES answers, Fairly credible: 4-5 YES answers, Less credible: 0-3 YES answers |  |  |

### Additional Analyses and Interpretations

We looked for evidence that super-spreading was more likely early in disease outbreaks. We interpreted the information collected with respect to whether persons identified as super-spreaders of novel coronaviruses ‘could be anyone’ or tend to fit a profile, such as being late middle age males or especially socially connected individuals. We were interested in whether super-spreading depended on certain types of people or the situations they are in. This information was only interpreted narratively but with respect to relevant literature or statistics such as the age distribution of persons in the country where the super-spreader lived.

We did not collect information on viral load carried by identified super-spreaders because this information is not systematically reported. However, we collected information to address the hypothesis that super-spreaders tend to be especially asymptomatic or especially severely ill. We compared the percentage of asymptomatic super-spreaders with published estimates of how many infections of each SARS/MERS/COVID are asymptomatic. Severity of disease was indicated by mortality outcomes. Survival rates for super-spreaders were calculated for each disease. The survival rates of patients who are super-spreaders were compared to publicly available survival rates of known MERS/SARS/COVID-19 cases.

## Results

### QUALITY ASSESSMENT

Altogether, 30% (36/119) of super-spreader individuals had epidemiologic descriptions that as a group, scored 6 in the quality assessment exercise (most credible rating). Proportionately, MERS super-spreaders had the highest number of “most credible” ratings (7/14 (50%)). Altogether, 28% (8/29) of SARS super-spreaders had epidemiologic evidence that was most credible, this figure was similar for COVID-19 super-spreaders (26% (21/76)). Figure 2 gives more detail about the distribution of quality scores for the individuals identified.

**Figure 1.**
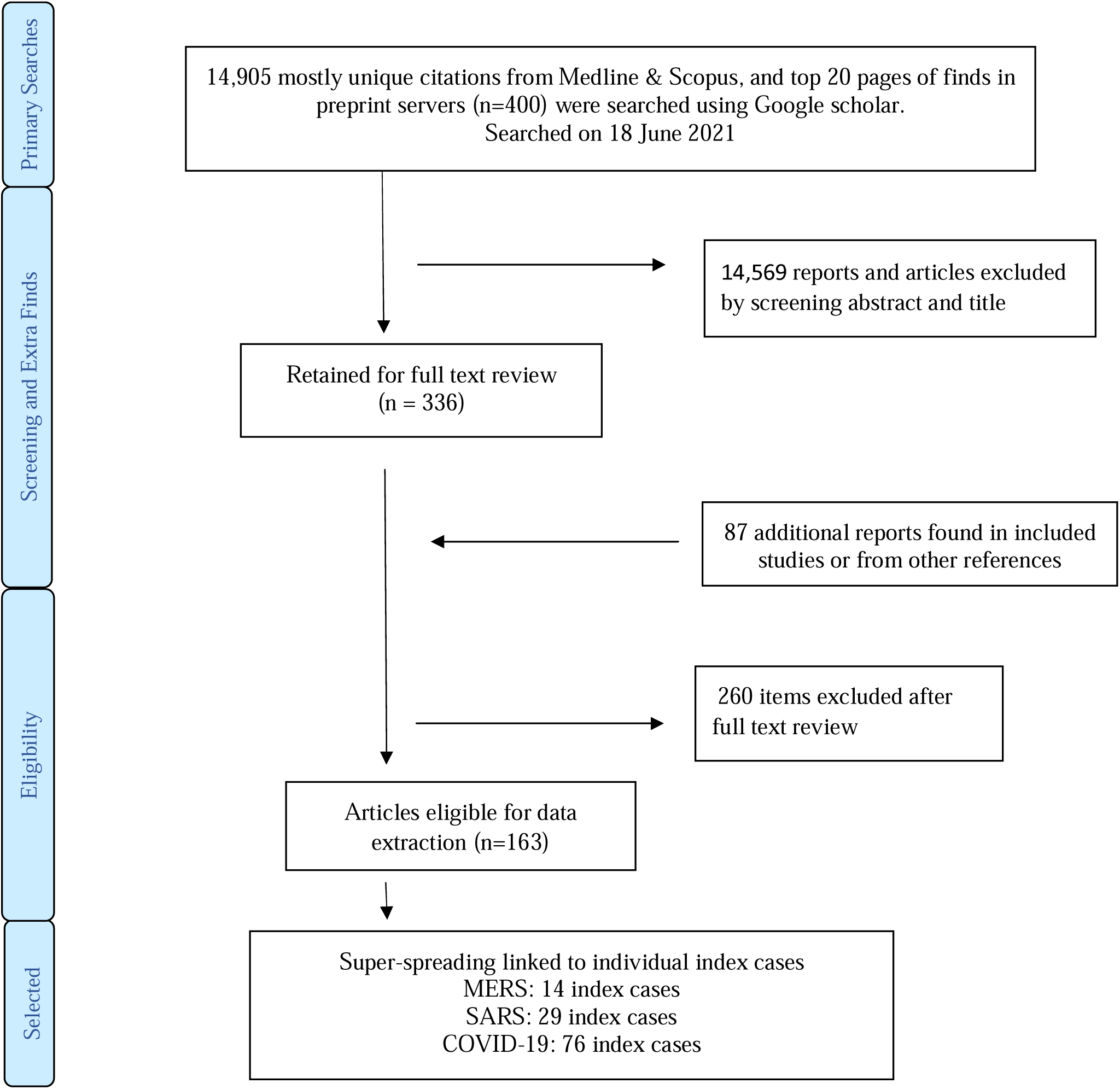
Report Selection Procedure

**Figure 2.**
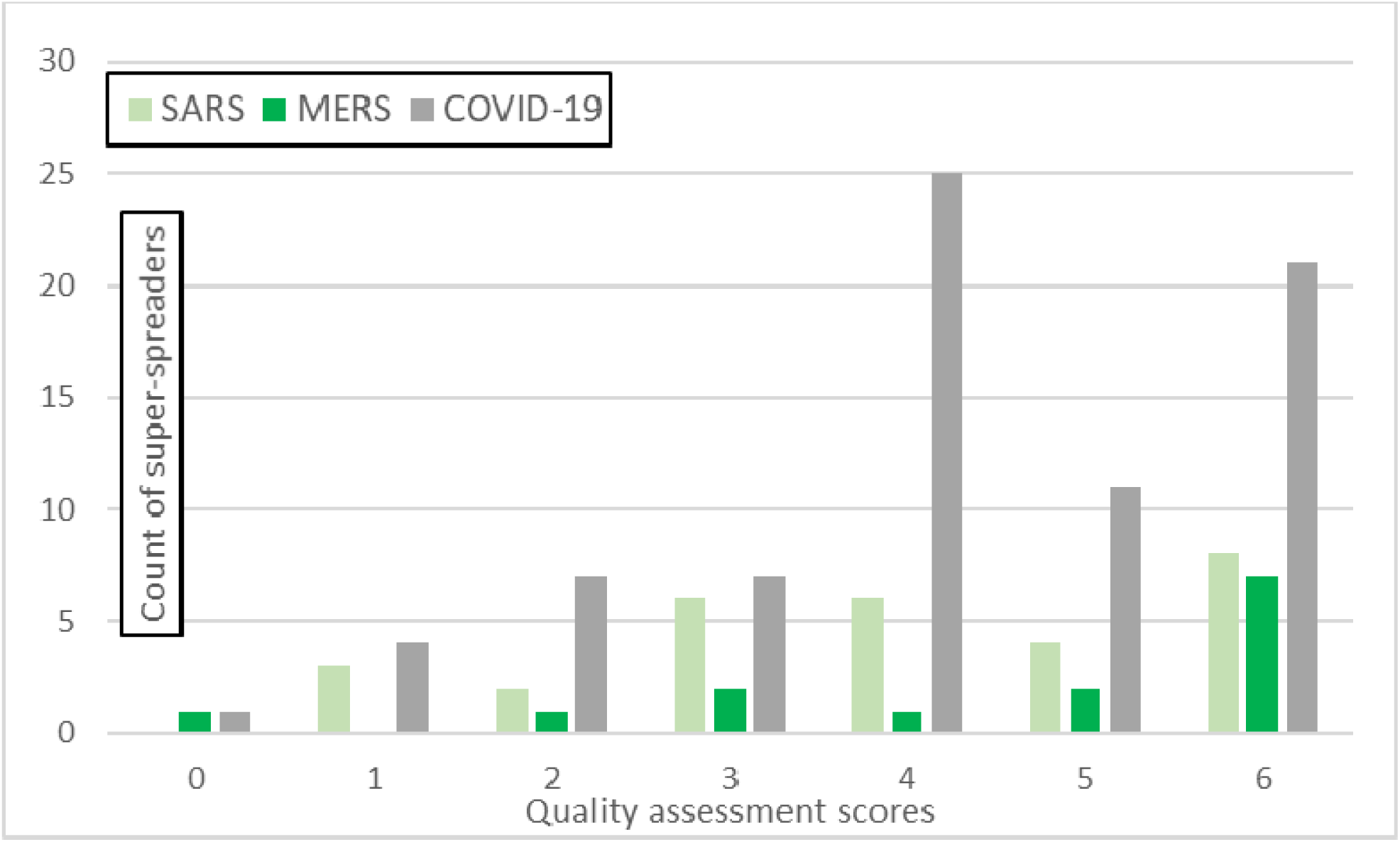
Quality assessment scores for epidemiologic information on nCoV super-spreaders.

### MERS

#### Counts

We provide a narrative summary because only 14 eligible MERS super-spreader individuals were identified. See Table S1 in Supplemental Material for MERS super-spreader details. Eight index cases were in Saudi Arabia, five in South Korea and one in the UAE. These transmissions were all in the period 2014-2017. The five South Korean super-spreaders were linked to the May-July 2015 MERS outbreak.

#### Activities when super-spreading and Settings

All MERS super-spreading was linked to time spent in clinical settings, usually as hospital inpatients who were admitted for their recognised respiratory illness, although day patients who attended clinic for other reasons (e.g., receiving dialysis) were also identified. Most secondary cases were other hospital patients and health care workers.

#### Age and sex

Index case age and sex was available for 12 individuals, who had median age 48 years (range 23-75 years). There were two females and 10 males.

#### Ethnicity and Occupation

Traits such as ethnicity and occupation were mostly not reported. Two super-spreaders were stated to be Korean, one was Yemeni, and another (patient in UAE) was described as ‘ex-patriate’. Occupation was only described for four MERS super-spreaders. In the Middle East, stated occupations were police storage room staff, camel butcher and factory worker. A hospital emergency department security room guard was a super-spreader in South Korea. The vast majority of secondary cases were themselves health care workers.

#### Secondary case counts

Estimated counts of secondary cases caused by these 14 index patients ranged in published reports (sometimes quite preliminary) from two, patient 16 in (Cowling *et al*. 2015) to 89, patient 14 in (Korea Centers for Disease & Prevention 2015). The largest counts of identified secondary cases and total persons in epidemiological clusters were in South Korea.

#### Symptom severity and outcomes

No MERS super-spreaders were described as asymptomatic; specific symptoms of severe illness were documented for nine super-spreaders among whom pneumonia was the single most common diagnosis (n=5). No information was given on survival or otherwise of two MERS super-spreaders. Of the remaining 12 super-spreaders, four survived to discharge and eight were recorded as deceased from MERS (mortality rate 67%). The crude case fatality rate for MERS has been reported as 34.8% (WHO 2017), so MERS super-spreaders may be more ill than most MERS cases.

#### Underlying conditions

Two of the MERS super-spreaders were investigated for comorbidities but reported to have no underlying health conditions; information was not provided for two others. Among the 10 super-spreaders with documented underlying conditions, a range of conditions (such as hypertension and kidney disease were mentioned): diabetes mellitus was the most commonly mentioned comorbidity (four of the 14 super-spreaders).

### SARS

Altogether, 29 eligible SARS super-spreader individuals were identified. Of these, 16 were in China, five in Singapore, three each in Canada and Hong Kong, and one each in Vietnam and Taiwan. All transmissions occurred in the period December 2002 to April 2003 (most were in March 2003). See Table S2 in Supplemental Material for summary extracted SARS super-spreader details.

#### Activities when super-spreading and settings

Almost all transmissions occurred while patients were in clinical settings (usually when they were admitted to hospital for treatment related to their SARS, but also when being treated as outpatients or visiting other inpatients). Some transmission in the community was deemed likely for five super-spreaders. On an aeroplane and other travel settings were identified as transmission locations for one patient each.

#### Age and sex

Index case sex was available for 27 individuals. There were seven females and 20 males. Index case age was available for 23 individuals who had median age 54 years (range 22-91 years, IQ range 42-70, 5^th^ percentile age 22 years, 95^th^ percentile 90 years).

#### Ethnicity and Occupation

Ethnicity or national origin was only reported for nine SARS super-spreaders: five were described as Chinese, one Malay, one Filipino, one Asian American and one of ‘non-Asian’ descent. There was no evidence that ethnic representation was different from ethnic distribution in the origin areas. Occupation was mostly not reported. Five individuals had occupational links to clinical settings: an ambulance driver, clinical professor, nurse, hospital laundry worker and family physician. There were two businesspersons, one seafood merchant and one vegetable seller. A patient aged 73 or 74 was described as retired. Similar to large MERS outbreaks, where occupational data were available for secondary cases, the secondary cases tended to mostly be health professionals themselves. Other occupations listed among the secondary cases were taxi drivers and market stall traders.

#### Secondary case counts

Estimated counts of secondary cases caused by these 29 index patients ranged from four to 51. Most cases were in the Far East (China, Hong Kong or Taiwan) plus Canada.

#### Symptom severity and outcomes

No SARS super-spreaders were described as asymptomatic. Information is available on the symptom severity for 22/29 SARS super-spreaders. Altogether, 18/22 are described as having fever; symptoms of respiratory illness were reported for 17/22. No information was given on mortality among 13 SARS super-spreaders. Of the remaining 16 patients, three survived to discharge and 13 were recorded as deceased from SARS (mortality rate 83%). SARS mortality rates are sex- and age-dependent, and were estimated at 26% for Chinese patients age ≥ 80 years (WHO 2003).

#### Underlying conditions

There was no information about possible underlying health conditions in 16 of the 29 SARS super-spreaders. Two were described as having unremarkable or ‘previously healthy’ histories. Underlying health conditions were reported for eleven patients. Eight of the 11 had cardiovascular conditions, six had forms of diabetes.

### COVID-19

In total, 76 eligible super-spreaders of COVID-19 were found. Of these, 38 were in east or southeast Asia, 15 were in Europe, 14 in North American and nine elsewhere in the world. The USA contributed 12 individuals to the database, the largest count from a single sovereign country. The vast majority (61 of 76) of their super-spreader events occurred before end of May 2020, so relatively early in the COVID-19 pandemic period. The largest count of super-spreaders found after 31 May 2020 were in the USA (7 super-spreader individuals described as index cases in the period June to August 2020). See Table S3 in Supplemental Material for summary of extracted COVID-19 super-spreader details.

#### Activities when super-spreading and Settings

Information on activities that were happening when super-spreading events happened was available for 56 individuals. In contrast to the MERS and SARS outbreaks, most super-spreading COVID-19 index patients were active in the community at the time that they were index patients, not clinical inpatients or receiving medical care. Just 12/56 of the super-spreaders were linked to clinical settings, either as patients or health care workers interacting with patients and colleagues. Other activities or settings linked to super-spreading were social/business in nature (n=22), religious worship (n=7), fitness or sport (n=5), education (n=4), public transport (n=3) and residential settings (one each of prison, summer camp and long term care facility).

#### Age and sex

Index case age was mostly unavailable; age data were most available in Asian countries. Approximate age information was provided for 28 individuals who had median age 56 years (range 18-85 years, IQ range 43-67, 5^th^ percentile age 21 years, 95^th^ percentile 85 years). From the 36 cases where sex was reported, there were 23 males and 13 females. All 13 super-spreaders whose sex was reported as female were from countries in east and southeast Asia (where males were equally represented, n=12). It is very possible that any sex imbalance for COVID-19 super-spreaders is an artefact of widespread lack of reporting index patient sex rather than evidence of underlying sex disparity in representation.

#### Ethnicity and Occupation

Ethnicity was rarely reported (only for six of 76 individuals). Where ethnicity was reported it was described by nationality typically, as “American” (n=3), “British” (n=2), and Korean. Occupation was also mostly not reported but was described for 26 of 76 individuals (34%). There were six health care workers, four fitness instructors, four school staff, three religious leaders, two retired individuals, two businesspersons, and one person each described as office worker, summer camp staff, sales representative, pilot and meat processing plant worker. One person may be plausibly inferred as a working as a church organist or piano player (Katelaris *et al*. 2021).

#### Symptom severity and outcomes

Symptom status was recorded for 50 individuals. Seven were described as asymptomatic at the time of super-spreading. Four of these persons subsequently developed symptoms during their detectable infection period (in Lam *et al*. 2020, Leung *et al*. 2020, Mahale *et al*. 2020, Yu *et al*. 2020,\ Cheng *et al*. 2021, Groves *et al*. 2021). Among our identified Covid super-spreaders, three were described as asymptomatic at all times during their infection, 13 as pre-symptomatic, and three as very mildly symptomatic. Other individuals had symptoms at the time of their super-spreading. No information on survival was available for 41 index cases. Three were reported to have died and 32 were reported to have survived. This suggests a case fatality rate for super-spreaders of about 8.8%, which is 3.27 times higher than all-age the COVID-19 case fatality rates suggested by pooled analysis of data available in the first 9 months of the COVID-19 pandemic (Ahammed *et al*. 2021).

#### Underlying conditions

Individual underlying health status was rarely reported for COVID-19 super-spreader individuals. It is likely that in some cases this lack of information means absence of underlying conditions, but we cannot be sure without explicit statement. Just four super-spreaders were reported to have underlying conditions, while one was reported to have had a previous “record of good health”.

The lack of specific age data on most super-spreader individuals resulted in small available datasets within any individual sovereign territory, making robust comparison with age distribution in each country untenable. The country that contributed the most specific age data most often was China. We therefore undertook a limited (China-only) comparison with the with age-distribution estimates within the Chinese population of 2003 (SARS) or 2020 (COVID-19), see Supplemental Figure S4. This limited comparison suggests that older individuals (age 40+) were over-represented for both SARS and COVID-19 compared with age-distribution estimates within the Chinese population of 2003 (SARS) or 2020 (COVID-19). These data suggest that individuals more prone to super-spreading are relatively older, at least age 40+.

## Discussion

Observable mortality rates for super-spreaders of any of the nCoV we studied were much higher than all-age mortality statistics for the same diseases. However, we note that mortality outcomes for COVID-19 index cases were especially poorly reported while in absolute terms, the count of COVID-19 super-spreaders who died was only 3 persons – so we may have an inaccurate picture of how often COVID-19 super-spreaders tend to die from the illness.

For SARS and MERS, super-spreaders tended to be quite ill. No asymptomatic super-spreaders were described for MERS and SARS, in spite of the prevalence of asymptomatic or mild infection among MERS cases being 26.9% (WHO 2017). This may be because for MERS and SARS only symptomatic persons were tested. Universal screening of health care contacts in 2003 found that 7.5-11% of persons with verified SARS-CoV infection were asymptomatic (Wilder-Smith *et al*. 2005, Nishiyama *et al*. 2008). This contrasts to COVID-19, where super-spreaders who were completely asymptomatic throughout their disease course, or who were presymptomatic at the time that they transmitted disease to many others, was quite commonly reported. That COVID-19 is likely to be most infectious very early in the disease course, before symptoms manifest (in contrast to SARS-CoV or MERS-CoV) was suggested by measurements of viral load in affected individuals (Cevik *et al*. 2021). At least one third of all SARS-CoV-2 infections were estimated to be asymptomatic in meta-analysis (Sah *et al*. 2021). This high prevalence of undetected infection has severely challenged pandemic control. We acknowledge that biases in ascertainment may have affected epidemiologic reports of symptom severity, especially for COVID-19. Individuals may have been motivated to incorrectly mis-describe their symptom status as asymptomatic when they had social contact, due to legal proscription against social contact for persons with COVID-19 symptoms in their jurisdictions at the time of their super-spreading.

It is noteworthy that few persons (n=2) under 20 years old were identified as super-spreaders for any of these novel coronaviruses (only for COVID-19). One of these persons was age 18 and the other was described as a teenage staff member; no younger children were described. There were many reports without a specific age, but in all cases the index case occupation or other social status (such as “father of the groom”) was sufficient to confirm that these persons with unspecified age were adults and not children. In addition, there were few super-spreaders identified to be in the 19-39 age category. This is an interesting result as younger persons tend to have much higher social contact rates than older individuals, which implies younger persons would be much more likely to become super-spreaders (Mossong *et al*. 2008). However, because COVID-19 is least likely to result in symptomatic illness among relatively young persons, and droplet spread of infectious respiratory disease tends to closely correlate with symptom severity (Bischoff *et al*. 2013), it is perhaps not surprising that we found relatively few super-spreaders under the age of 40 for any of the novel coronaviruses. Most of the super-spreading events found in our review happened when widespread social distancing measures were not in place in the relevant jurisdictions and social contact rates were therefore relatively ‘normal’. This suggests that social-distancing policies were unlikely to have distorted social contact patterns that vary by age group. If there is a lower risk that relatively younger persons are less likely to be super-spreaders, this knowledge has bearing on whether social distancing or education provision for these age cohorts can be managed differently than for other age groups in any future nCoV outbreak.

We tried to identify super-spreaders who were identified as having caused at least nine secondary infections by any one report. However, the counts of secondary infections from individual index cases varied in the epidemiological reports. We did not aspire to evaluate which reports were most accurate (in absence of all primary epidemiological data). Therefore the counts of secondary cases range from values below our eligible minimum (i.e., were sometimes below nine).

We did not collect information on posited transmission pathway (e.g., droplet, aerosol or airborne) and cannot report on this definitively. That super-spreading might be especially linked to aerosol spread was hypothesised early on for COVID-19 (Günther *et al*. 2020). Among the super-spreader individuals we identified, an aerosol transmission pathway was especially well described in (Sugimoto & Kohama 2020) and apparent airborne transmission as the only plausible main transmission pathway was documented in (Cheng *et al*. 2021). We note that the definition of what is “airborne transmission” has been redefined by the COVID-19 pandemic (Tang *et al*. 2021) and this categorisation seems likely to continue to evolve, making it unfeasible for our review to contribute certain data at this time. We can note that most of the activities that super-spreader individuals in our database were engaged in at the time that they were index cases, where information was available, suggest that a droplet-transmission pathway was just as likely as air, aerosol or surface transmission pathways.

There was insufficient information about occupation to link any specific line/s of work to likely super-spreading. However, we note that health care workers were especially likely to be secondary cases for MERS and SARS. There was very limited information about patient ethnicity and frequently this was reported as nationality. Based upon the limited information available there was no indication that super-spreaders had unusual ethnicity compared to majority of population in the areas where they lived.

Overall there appears to be some differences between super-spreaders with MERS and SARS compared to COVID-19. The definition of a super-spreader is complex, as often it is unclear as to how many people have been infected from a single source, and for COVID-19, where the disease has high community prevalence this is particularly difficult. For all three diseases there is little evidence for younger people to be super-spreaders, however this may reflect the timing of this study, in particular for COVID-19. However, based on our study it appears that for MERS and SARS, super-spreaders are more likely to be patients who have more severe symptoms, while for COVID-19 asymptomatic sufferers appear to be important. These findings suggest the value of high awareness of asymptomatic COVID-19 infection as a potential for super-spreading, given the high level of community transmission that has existed in some areas.

## Data Availability

Extracted dataset used for analysis is appended; the extracted data were already published.

## Limitations

Ascertainment bias affected our efforts. Universal screening of all MERS and SARS contacts rarely happened, in contrast to much broader testing of all COVID-19 contacts. Therefore, it is unsurprising that we detected and collected data about many more COVID-19 super-spreader individuals. However, even when super-spreading individuals are detected within a surveillance system a decision is then made whether to publish as a case report or academic publication and it is unknown what biases this introduces. Case reports may be more likely for diseases perceived to be rare and hence the proportionally greater number for MERS and SARS may be unsurprising. It is also possible that super-spreaders who are considered unusual or have already generated publicity are preferentially published.

In order to systematically collect data about super-spreading individuals we had to create consistent criteria to identify them. This means inevitably that slightly different criteria would have identified a different group of persons and dataset to describe. However, we do not believe that such alternative thresholds would have greatly changed the impression we gained about what kinds of people tend to be nCoV super-spreaders.

We could have used a different secondary case threshold (to more or less than nine secondary cases). We chose a seemingly high threshold for secondary cases to minimise ambiguity about whether our definition of ‘super-spreading’ was in excess of typical number of secondary cases generated. When extracting data from individual publications the level of information presented varied greatly between reports. For example gender and age was not always reported and more systematic reporting would be beneficial moving forward. It is unclear whether these gaps introduced biases into our results.

Our searches were updated to mid June 2021; data about superspreading linked to the Delta variant of COVID-19 were unlikely to be included while the Omicron or later variants could not be included (because we collected data before Omicron emerged). COVID-19 is fast moving and our findings can only be relevant to the time period investigated. We have reported attributes, such as symptom severity, as they were described in the original reports. It may be that some attributes (such as symptom severity) have not been reported faithfully

## Conclusion

One of the key questions posed by Stein (2011) is what makes individuals into super-spreaders. We demonstrate that this key question is hard to answer due to the limited number of published studies on super-spreader individuals, but more importantly inconsistencies in the available details about index cases. More outbreak reports should be fully published with suitably anonymised descriptions of index and secondary patients. Since early 2020, detailed contact tracing databases have been assembled in many jurisdictions to facilitate COVID-19 control: these may form a rich resource for identifying super-spreaders and super-spreading too, but they need to be made available with relevant case demographic and outcome information. Should stronger data emerge, they could be incorporated into disease modelling or be used to guide intensive contact tracing in outbreak situations. For all three diseases we found that males and persons age 40+ were the most likely to be identified as super-spreaders of novel coronavirus diseases MERS, SARS and COVID-19, while persons under age 18 years were unlikely to be identified as super-spreaders. However, super-spreader characteristics varied between the coronaviruses. Most super-spreading from MERS or SARS was observed in clinical environments, while COVID-19 super-spreading happened predominantly in community settings. Generally, MERS or SARS super-spreaders were highly symptomatic and had poor disease outcomes and underlying health conditions. In contrast, many individuals who super-spread COVID-19 were observed to have mild or no symptoms at the time of their high transmission rate, and where survival status was documented for persons who had tested positive for SARS-CoV-2, the vast majority survived the infectious period.

## Supplemental Material

### Tables S1-S3 and Figure S4

**NOTES for all tables:** Quality scores are based on count of ‘Yes’ answers to quality checklist shown as Table 1. #secondaries means the number of dir ect secondary cases caused by the specified index case, according to the group of related articles that describe this index case, hence the answer can be a range if multiple sources have different estimates for the number of secondary cases linked to the index patient.

**Table S1.** Traits of MERS super-spreader individuals

| Name, age, sex | Dates/country | Activities/setting | Estimated #secondaries | symptoms & outcome | Underlying condition | Quality score | Key Source(s) |
| --- | --- | --- | --- | --- | --- | --- | --- |
| Pt III-A<br>45, M | 2014, UAE | patient in ED then ICU | 11 | Breathing difficulties.<br>Died | No information | 5 | (Hunter *et al.* 2016) |
| Pt1,<br>53, M | 2014,<br>Saudi Arabia | While receiving kidney dialysis, clinic | 14 | Symptoms not reported.<br>Died | In receipt of regular dialysis | 5 | (Assiri *et al.* 2016) |
| Index,<br>no info | 2015,<br>Saudi Arabia | patient in ED | 10 | No information | No information | 3 | (Saad *et al.* 2014) |
| Case 02,<br>56, M | 2015,<br>Saudi Arabia | Hospital ED and inpatient | 8-9 | Symptoms not reported.<br>Died | Diabetes, hypertension, hypothyroidism, coronary artery disease | 4 | (Alenazi *et al.* 2017, El Bushra *et al.* 2017, Barry *et al.* 2020) |
| Pt 1664<br>~48, F | 2016,<br>Saudi Arabia | Hospital inpatient | 17-23 | Symptoms not reported.<br>Died | Unspecified comorbidities | 2 | (Adegboye & Elfaki 2018) |
| Pt1,<br>68, M | 2015, S.Korea | Seeking treatment in ED, later as hospital inpatient | 19-38 | pneumonia, severe cough, hospitalised for months. Survived | hypertension | 6 | (Cowling *et al.* 2015, Kim *et al.* 2015, Korea Centers for Disease & Prevention 2015, Park *et al.* 2016) |
| Pt14, 35, M | 2015, S.Korea | Seeking treatment in ED, later as hospital inpatient | 73-93 | pneumonia, severe cough, respiratory failure, diarrhea. Survived | Ascertained but none known | 6 | (Cowling *et al.* 2015, Kim *et al.* 2015, Korea Centers for Disease & Prevention 2015, Park *et al.* 2016) |
| Pt15, 35, M | 2015, S.Korea | As hospital inpatient | 4-9 | pneumonia without severe cough. Survived. | Liver failure | 3 | (Nishiura *et al.* 2016, Kim *et al.* 2017) |
| Pt16, 41, M | 2015, S.Korea | As hospital inpatient | 21-25 | pneumonia, severe cough. Survived | pancreatitis | 6 | (Korea Centers for Disease & Prevention 2015, Choe *et al.* 2020) |
| Pt76, 75, F | 2015, S.Korea | As hospital inpatient | 10-12 | pneumonia, slight cough. Died. | multiple myeloma; diabetes mellitus | 6 | (Korea Centers for Disease & Prevention 2015, Choe *et al.* 2020) |
| Case 5, 65, M | 2017, Saudi Arabia | Seeking treatment in ED, later as hospital inpatient | 9-11 | Symptoms not reported. Died | Diabetes Mellitus, Hypertension | 6 | (Alanazi *et al.* 2019) |
| Index A, ~47, M | 2017, Saudi Arabia | Seeking treatment in ED, later as hospital inpatient | 16-19 | Cough, Shortness of breath, chest pain. No fever. History of Diarrhea prior to admission. Died | Diabetes Mellitus, Hypertension, Chronic Kidney Disease | 6 | (Alanazi *et al.* 2019) |
| Index B, 23, M | 2017, Saudi Arabia | Seeking treatment in ED, later as hospital inpatient | 9 | Fever, cough, rhinorrhea. Died | Ascertained but none known | 6 | (Alanazi *et al.* 2019) |
| Index, no info | 2017, Saudi Arabia | While receiving dialysis and within household | Up to 10 | No information | Was receiving dialysis | 0 | (WHO 2017) |

**Table S2.**
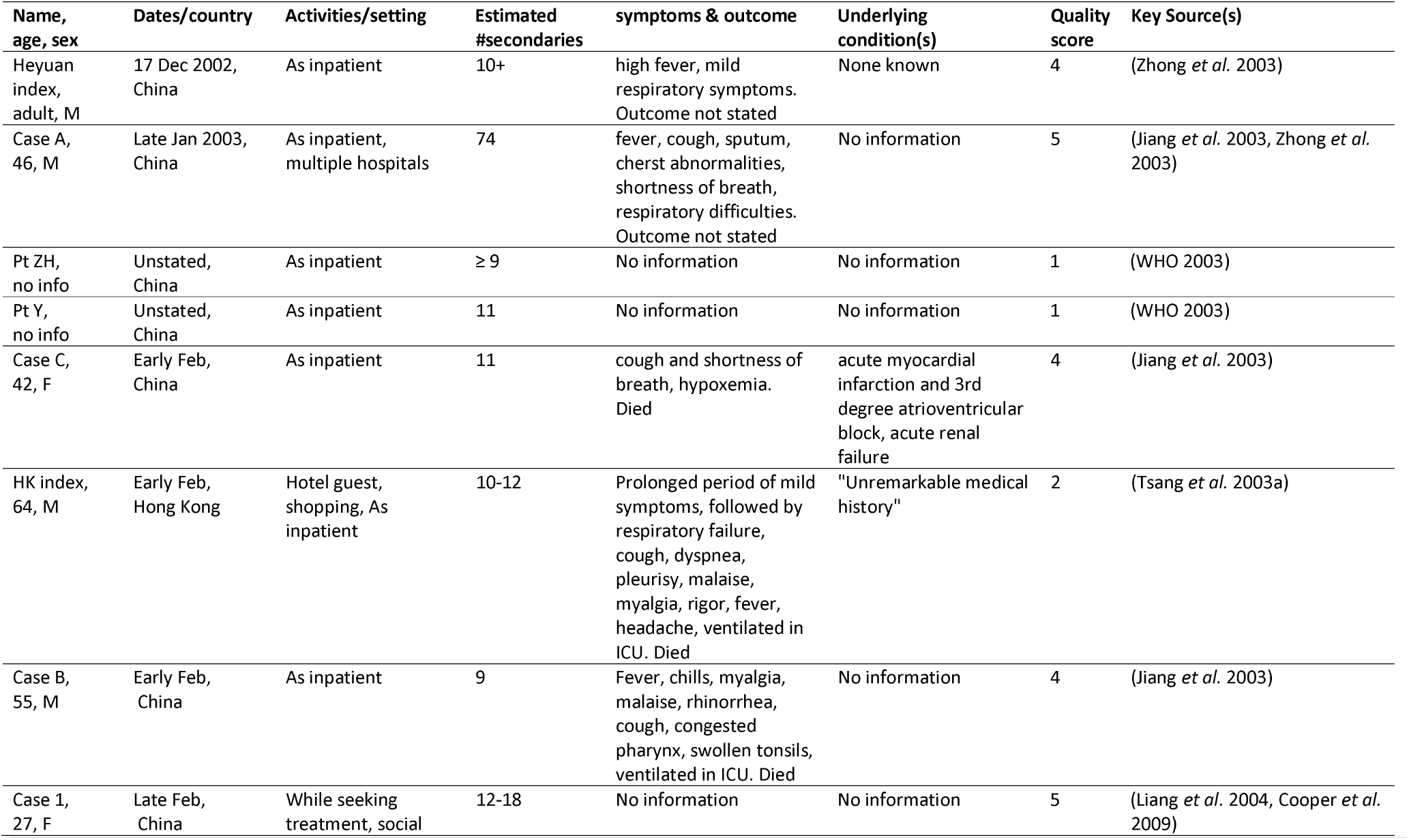
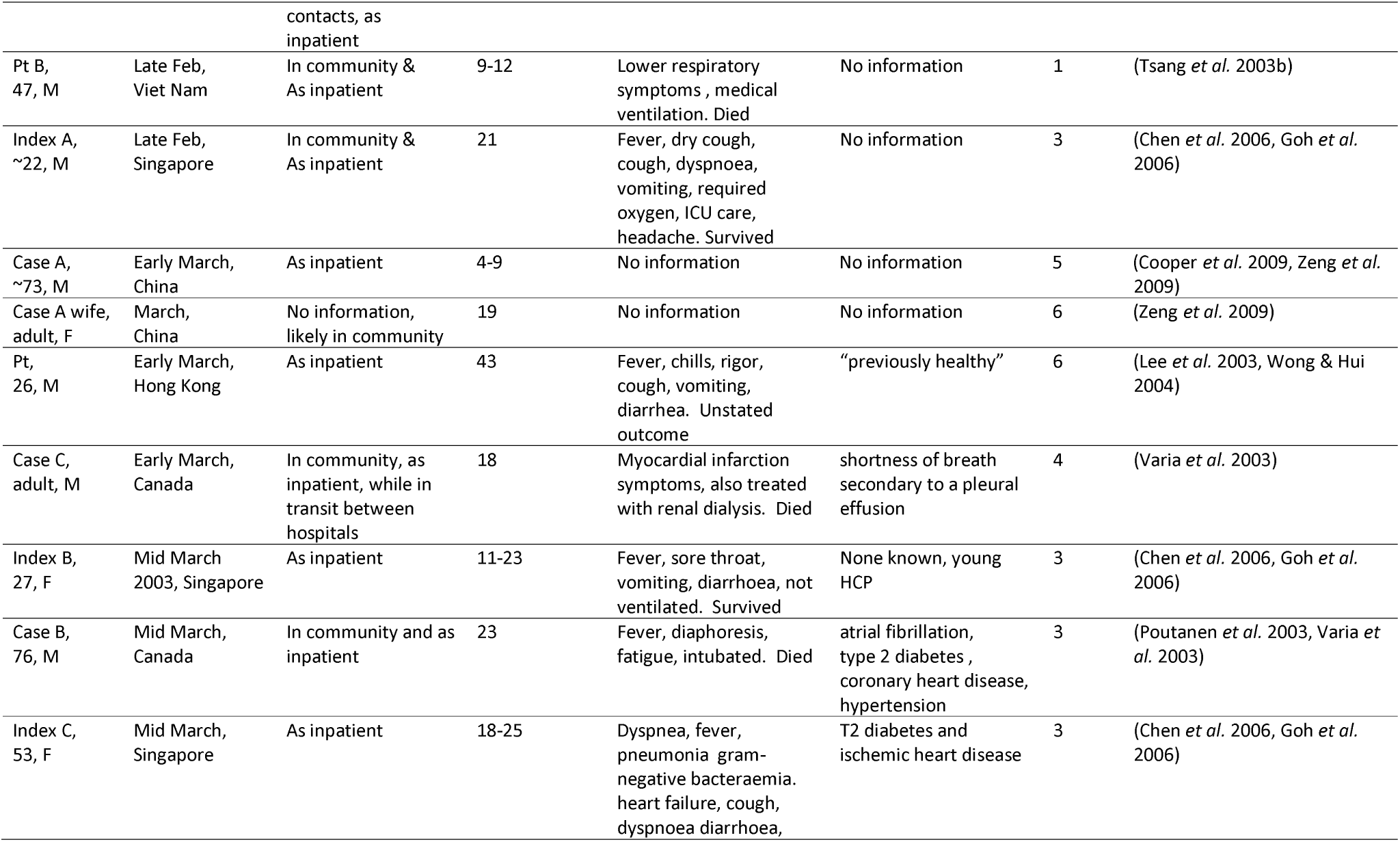
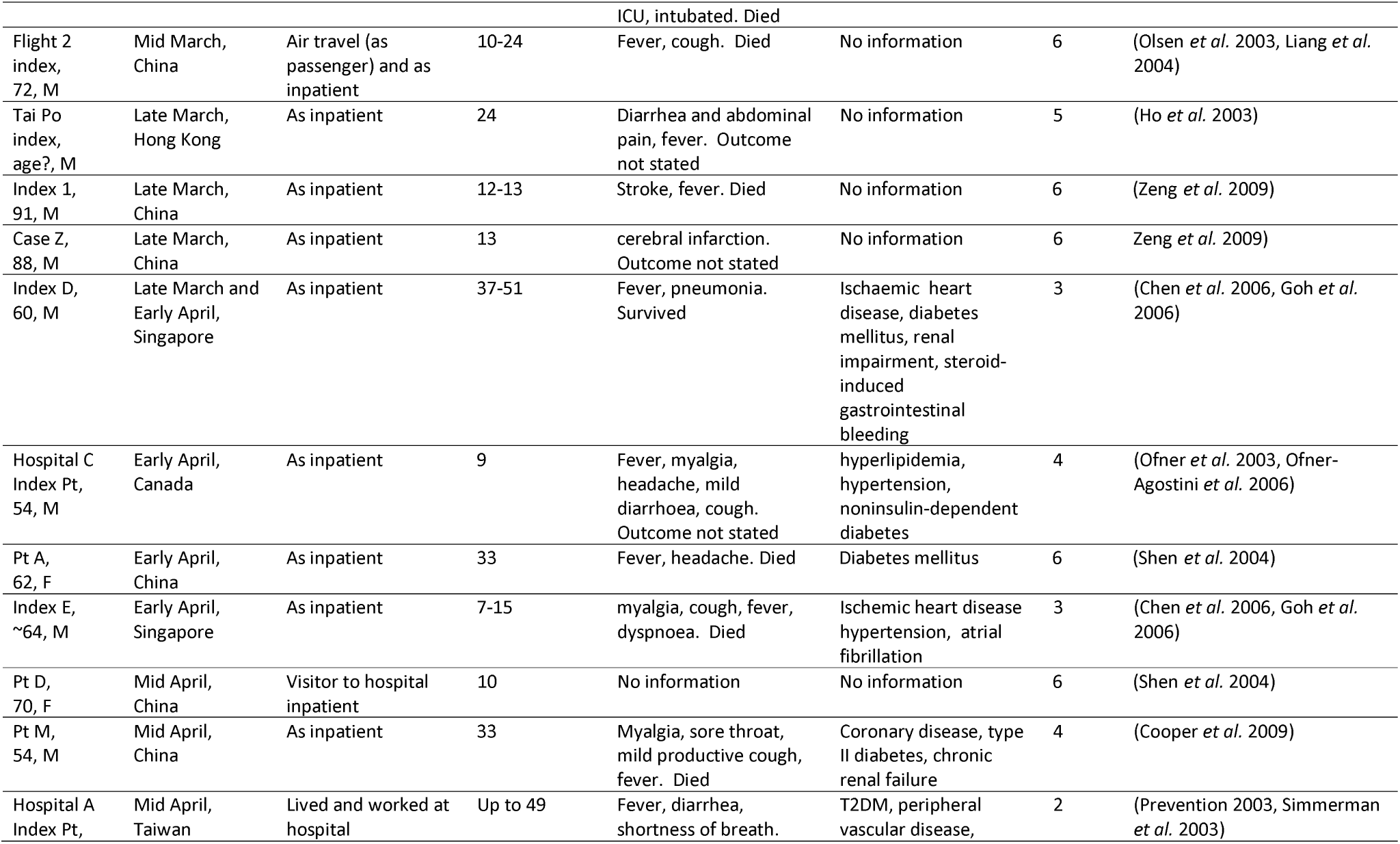
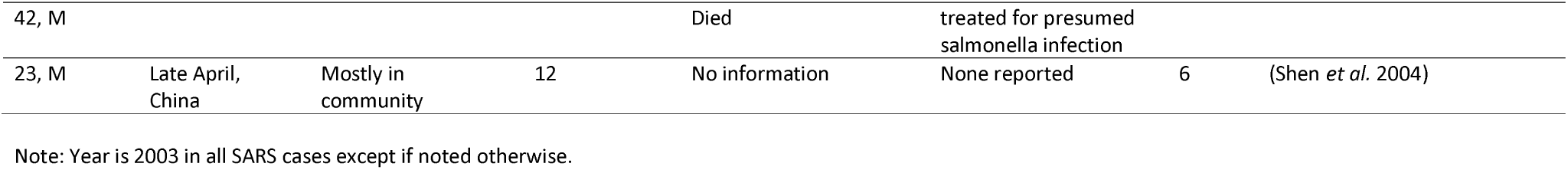
Traits of SARS super-spreader individuals

**Table S3.**
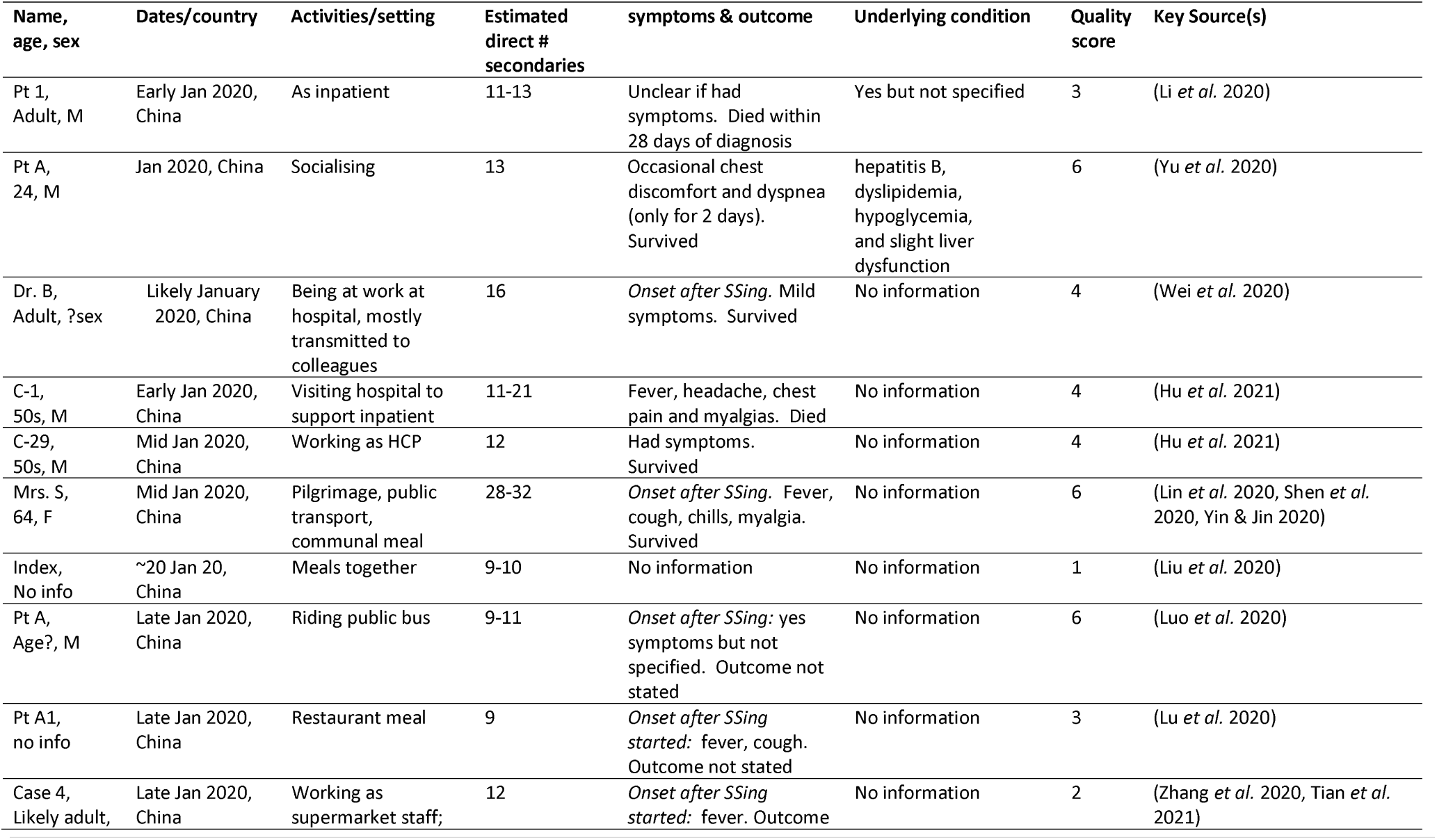
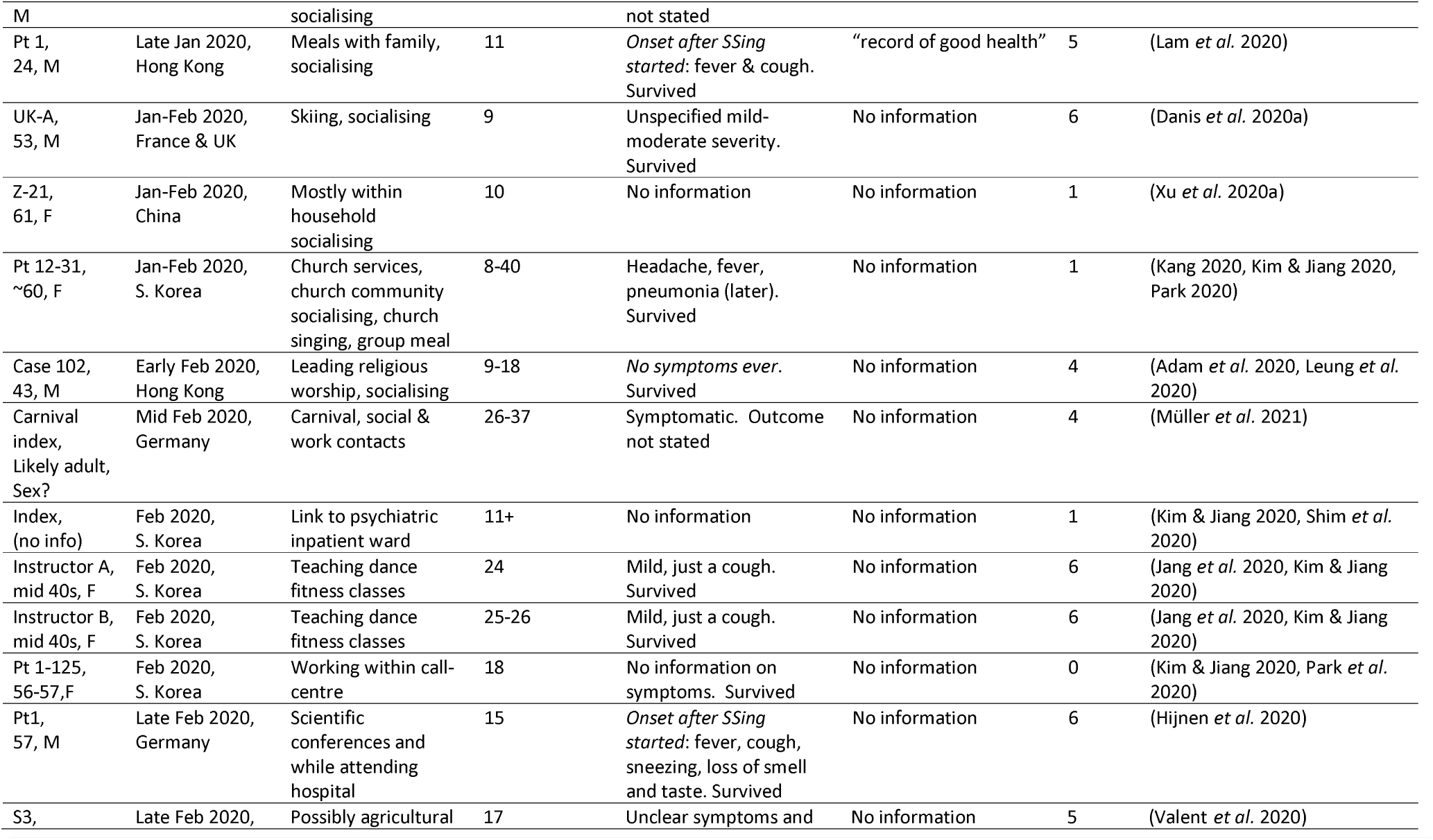
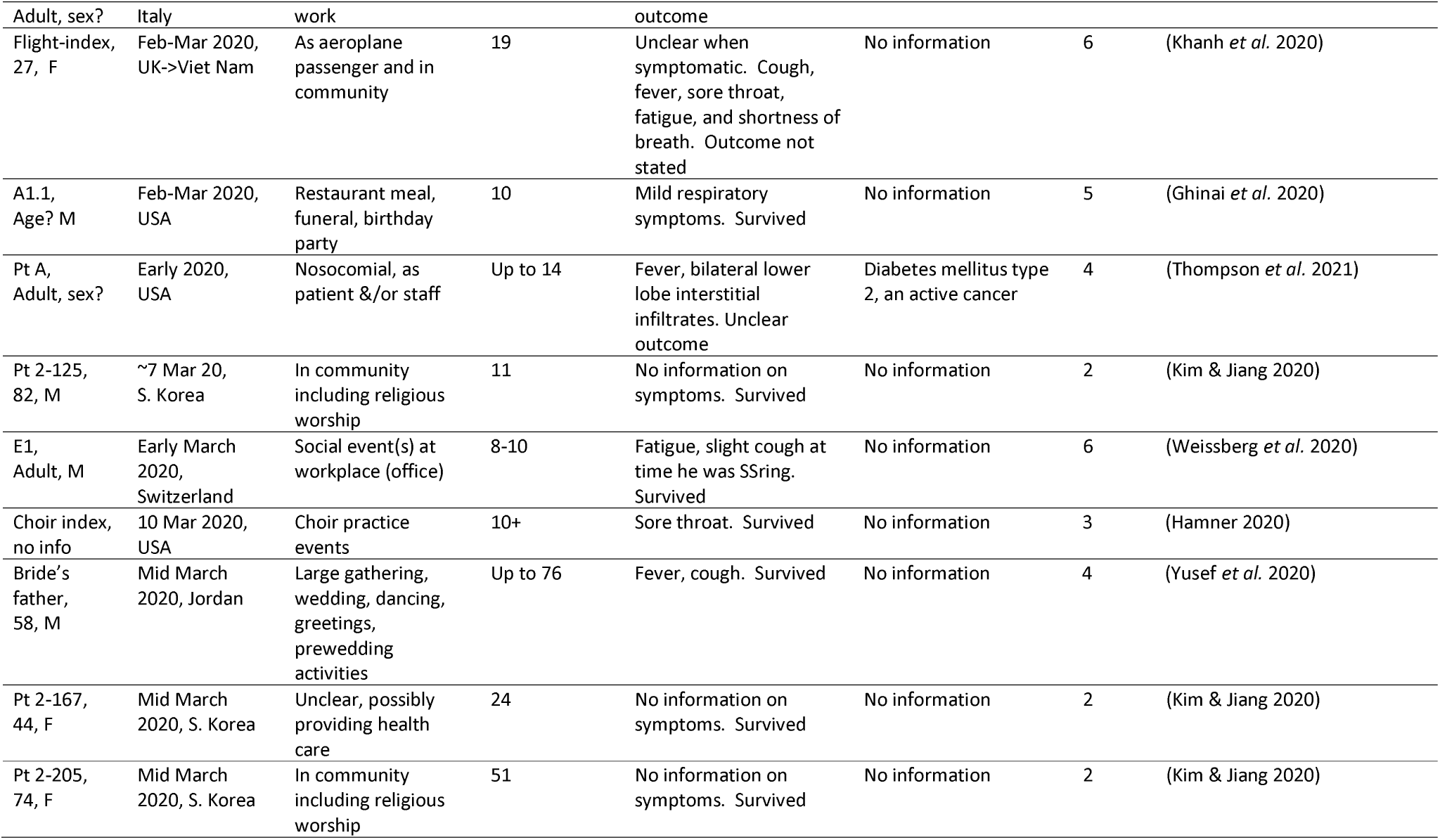
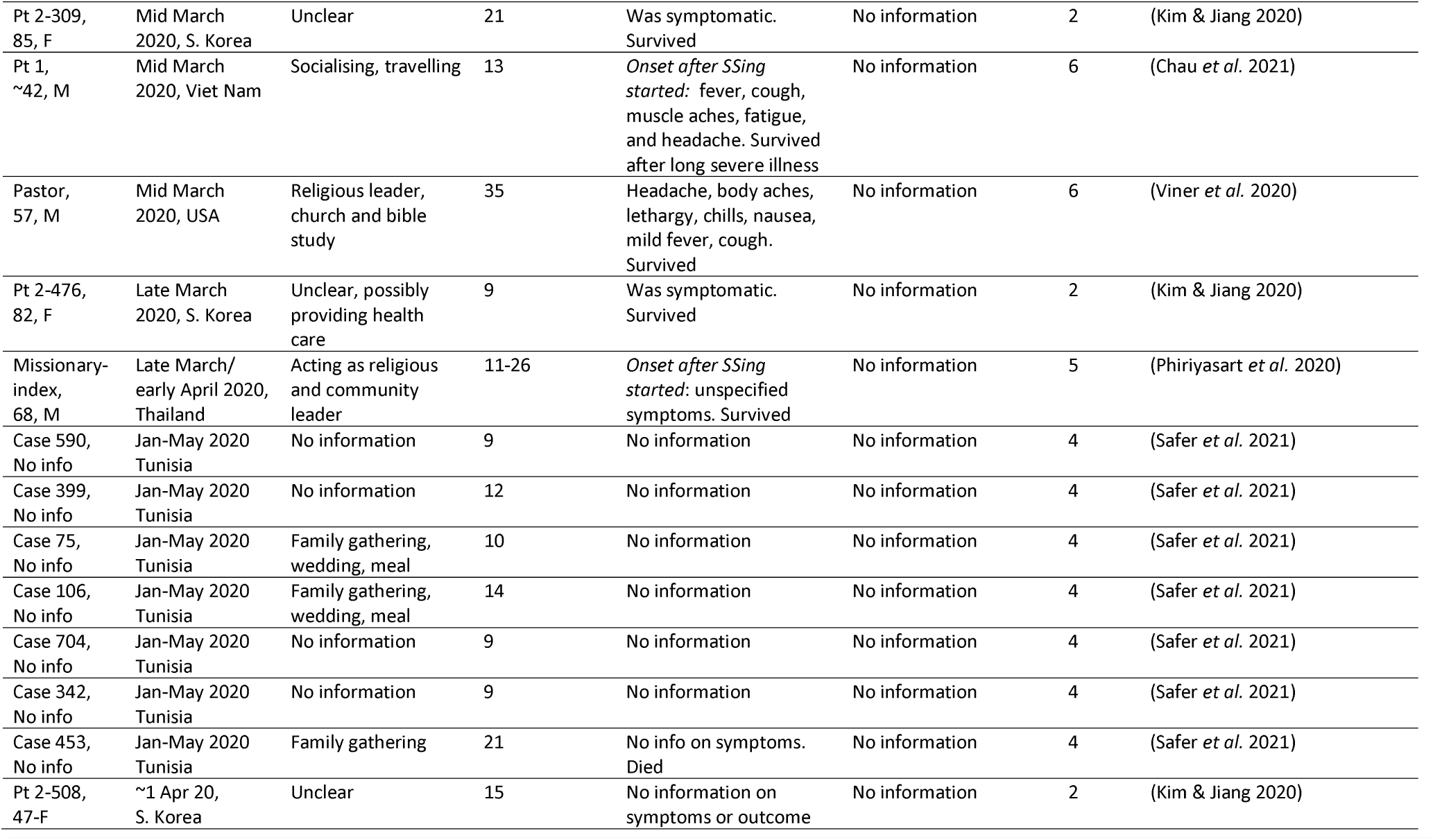
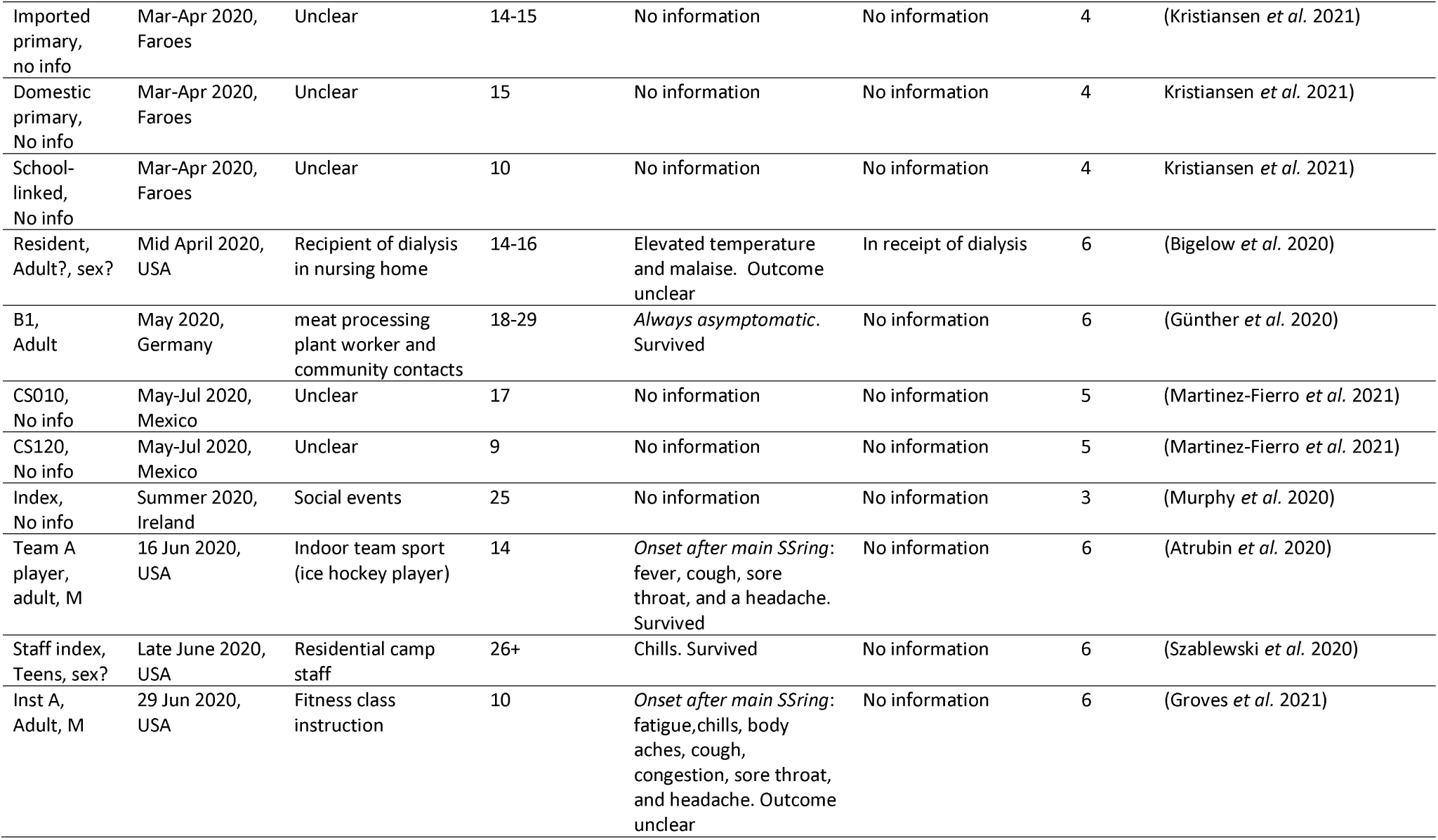
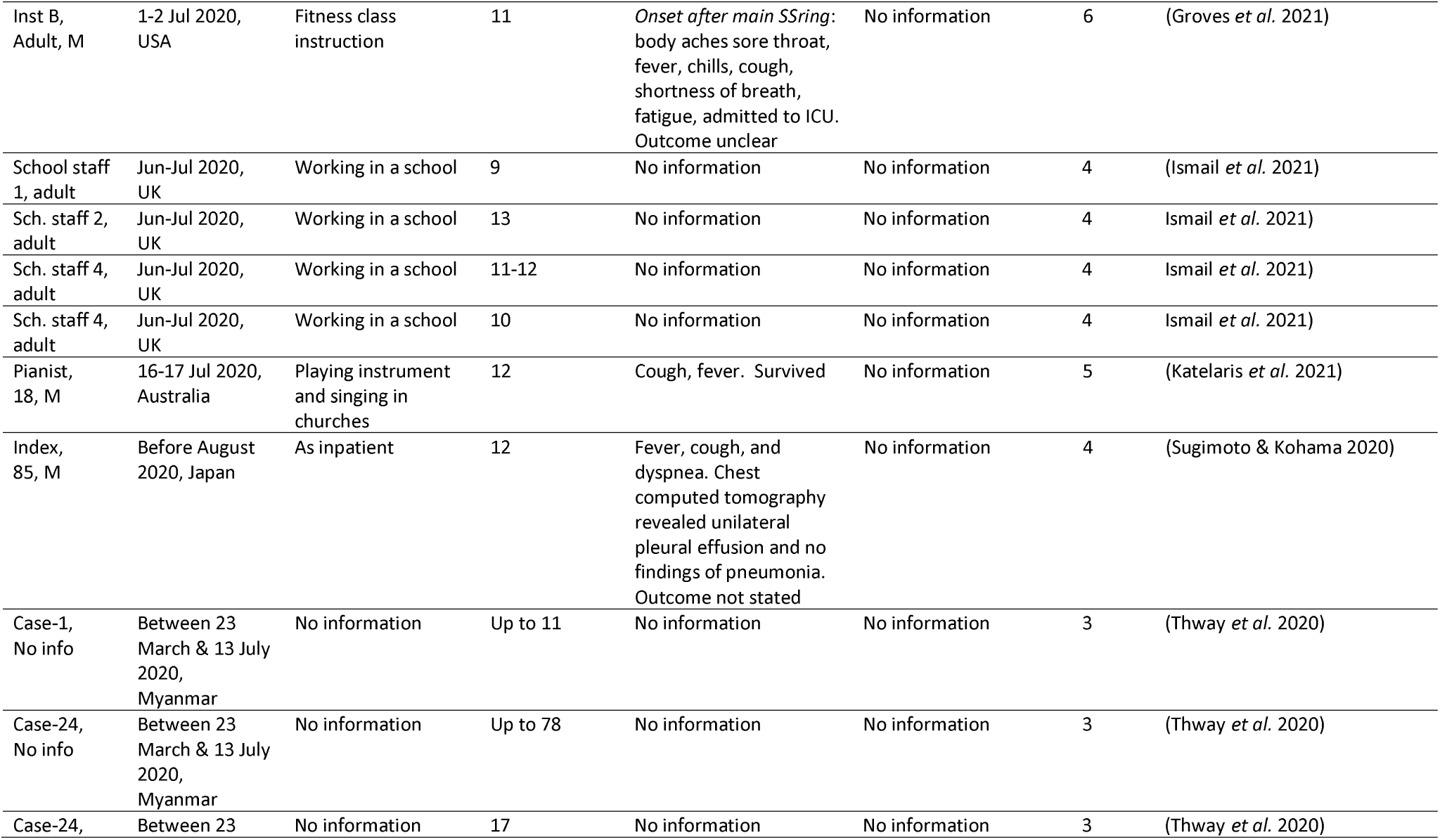
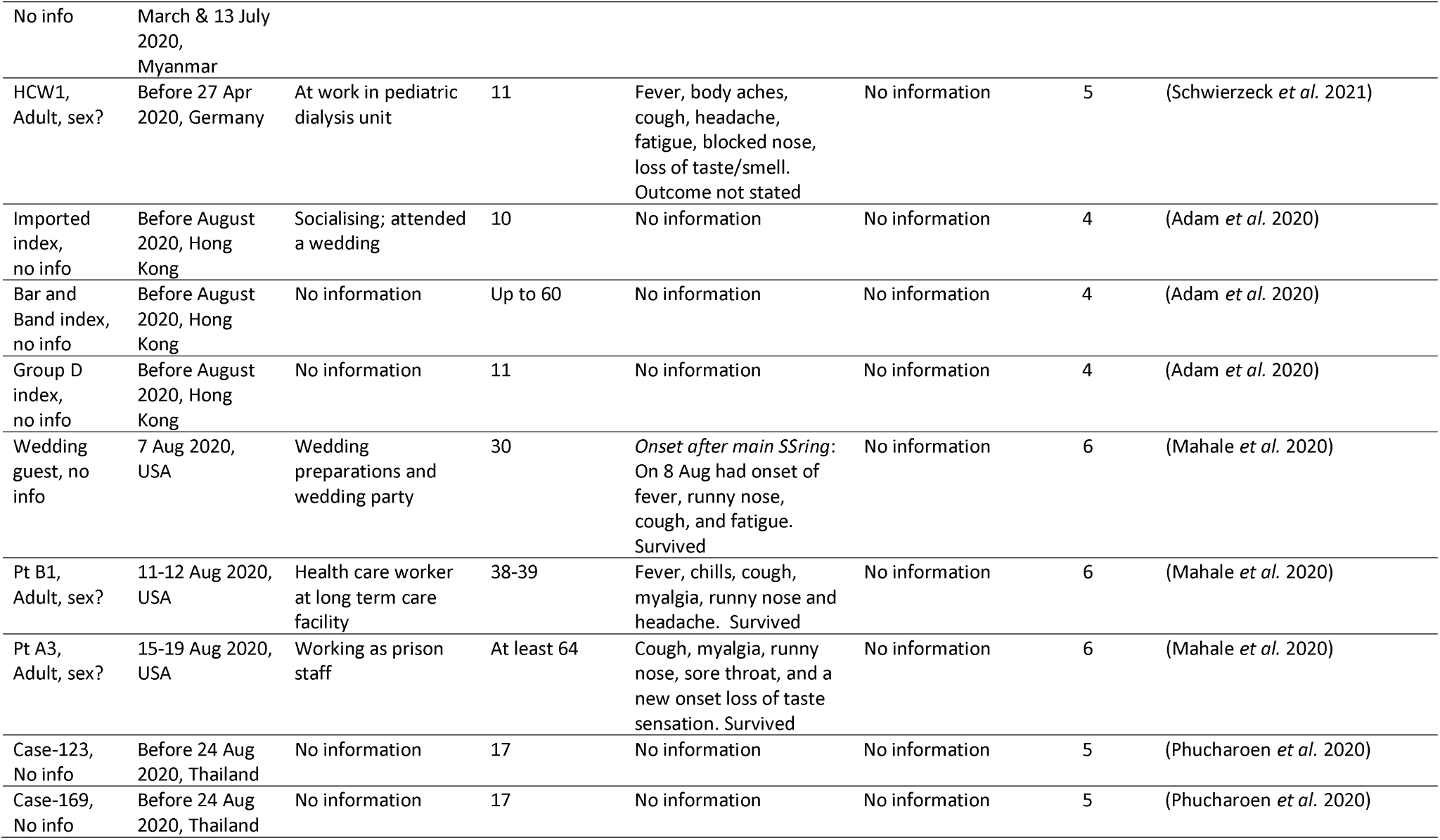
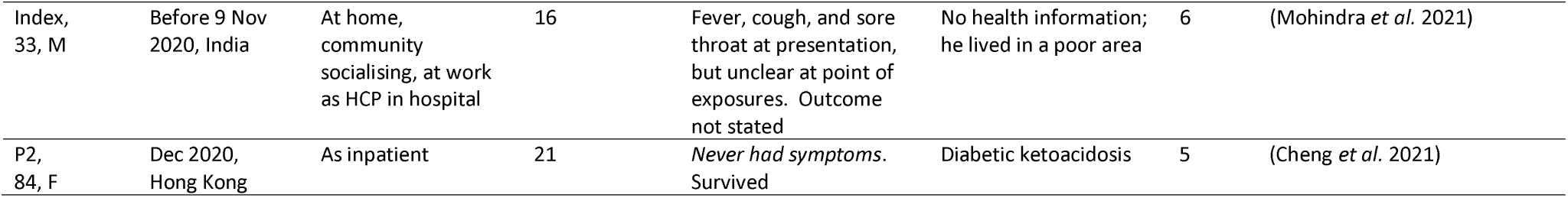
Traits of COVID-19 super-spreader individuals

**Figure S4.**
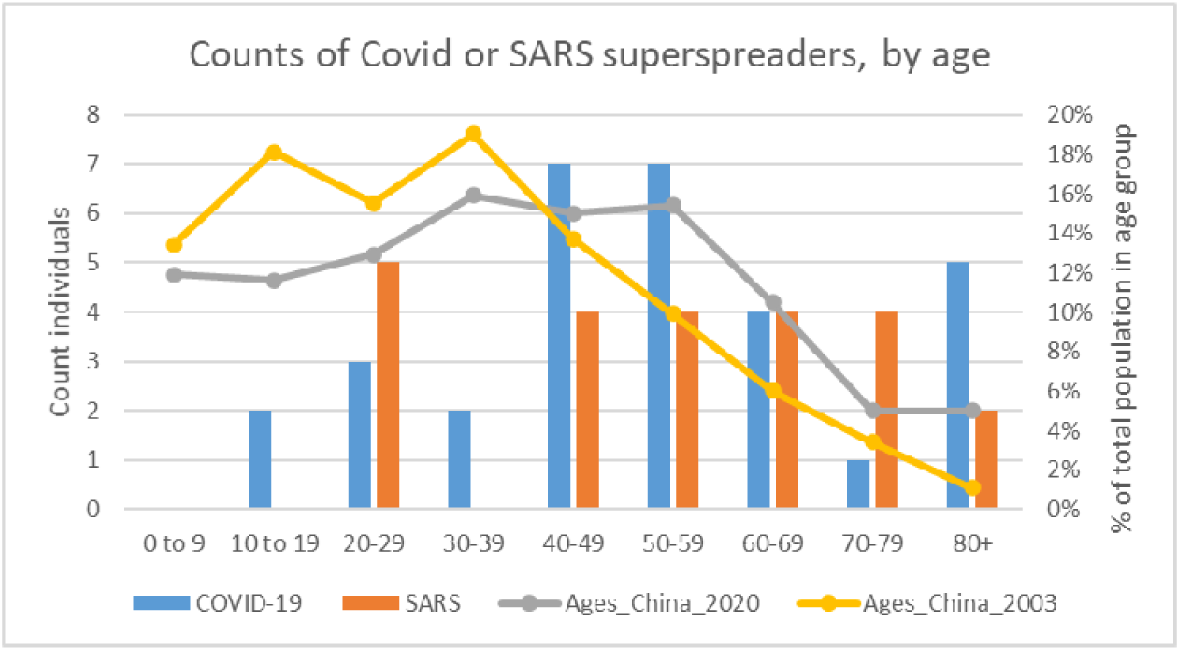
Age distribution of super-spreader cases compared to concurrent proportion of population in each age group in China. ***Note***: All super-spreader individuals are counted here but the population proportions in each age group are only for China in 2003 and 2020 because China contributed most of the age-specific data for either SARS or COVID super-spreaders Detailed breakdown or comparison for each contributing country is not informative when ages were generally unavailable outside of China. *Chinese Population distribution source*: https://www.populationpyramid.net/china.

## Notes

**Conflict of interest** The authors declare that we have no conflict of interest.

### Competing Interest Statement

The authors have declared no competing interest.

### Funding Statement

This study was funded by the National Institute for Health Research Health Protection Research Unit (NIHR HPRU) in Emergency Preparedness and Response at Kings College London in partnership with the UK Health Security Agency (UK HSA) and collaboration with the University of East Anglia. The views expressed are those of the author(s) and not necessarily those of the NHS, the NIHR, UEA, the Department of Health or UK HSA.

### Author Declarations

Extracted dataset used for analysis is appended; the extracted data were already published.

## References

Ahammed, T., A. Anjum, M. M. Rahman, N. Haider, R. Kock and M. J. Uddin (2021). “Estimation of novel coronavirus (COVID-19) reproduction number and case fatality rate: A systematic review and meta-analysis.” Health Science Reports 4(2): e274.

Al-Tawfiq, J. A. and A. J. Rodriguez-Morales (2020). “Super-spreading events and contribution to transmission of MERS, SARS, and SARS-CoV-2 (COVID-19).” Journal of Hospital Infection 105(2): 111–112.

Bischoff, W. E., K. Swett, I. Leng and T. R. Peters (2013). “Exposure to influenza virus aerosols during routine patient care.” The Journal of Infectious Diseases 207(7): 1037–1046.

Cevik, M., M. Tate, O. Lloyd, A. E. Maraolo, J. Schafers and A. Ho (2021). “SARS-CoV-2, SARS-CoV, and MERS-CoV viral load dynamics, duration of viral shedding, and infectiousness: a systematic review and meta-analysis.” The Lancet Microbe 2: e13–22.

Cheng, V. C.-C., K. S.-C. Fung, G. K.-H. Siu, S.-C. Wong, L. S.-K. Cheng, M.-S. Wong, L.-K. Lee, W.-M. Chan, K.-Y. Chau and J. S.-L. Leung (2021). “Nosocomial Outbreak of Coronavirus Disease 2019 by Possible Airborne Transmission Leading to a Superspreading Event.” Clinical Infectious Diseases 73(6): e1356–e1364.

Choi, S., E. Jung, B. Choi, Y. Hur and M. Ki (2018). “High reproduction number of Middle East respiratory syndrome coronavirus in nosocomial outbreaks: mathematical modelling in Saudi Arabia and South Korea.” Journal of Hospital Infection 99(2): 162–168.

Cowling, B. J., M. Park, V. J. Fang, P. Wu, G. M. Leung and J. T. Wu (2015). “Preliminary epidemiologic assessment of MERS-CoV outbreak in South Korea, May–June 2015.” Euro surveillance: bulletin Europeen sur les maladies transmissibles= European communicable disease bulletin 20(25).

Danis, K., O. Epaulard, T. Bénet, A. Gaymard, S. Campoy, E. Bothelo-Nevers, M. Bouscambert-Duchamp, G. Spaccaferri, F. Ader and A. Mailles (2020). “Cluster of coronavirus disease 2019 (Covid-19) in the French Alps, 2020.” Clinical Infectious Diseases(71).

Frieden, T. R. and C. T. Lee (2020). “Identifying and interrupting superspreading events—implications for control of severe acute respiratory syndrome coronavirus 2.” Emerging Infectious Diseases 26(6): 1059–1066.

Grey, H. (2020, May 27 2020). “Just One Person Can Transmit COVID-19 to Dozens in a Few Hours.” from https://www.healthline.com/health-news/how-one-person-can-transmit-covid19-to-dozens-in-just-a-few-hours.

Groves, L. M., L. Usagawa, J. Elm, E. Low, A. Manuzak, J. Quint, K. E. Center, A. M. Buff and S. K. Kemble (2021). “Community transmission of SARS-CoV-2 at three fitness facilities—Hawaii, June–July 2020.” Morbidity and Mortality Weekly Report 70(9): 316.

Günther, T., M. Czech-Sioli, D. Indenbirken, A. Robitaille, P. Tenhaken, M. Exner, M. Ottinger, N. Fischer, A. Grundhoff and M. M. Brinkmann (2020). “SARS-CoV-2 outbreak investigation in a German meat processing plant.” EMBO Molecular Medicine 12(12): e13296.

Kain, M. P., M. L. Childs, A. D. Becker and E. A. Mordecai (2021). “Chopping the tail: How preventing superspreading can help to maintain COVID-19 control.” Epidemics 34: 100430.

Kang, C. K., K.-H. Song, P. G. Choe, W. B. Park, J. H. Bang, E. S. Kim, S. W. Park, H. B. Kim, N. J. Kim and S.-i. Cho (2017). “Clinical and epidemiologic characteristics of spreaders of Middle East respiratory syndrome coronavirus during the 2015 outbreak in Korea.” Journal of Korean Medical Science 32(5): 744–749.

Kasulis, K. (2020). “‘Patient 31’ and South Korea’s sudden spike in coronavirus cases.” Al Jazeera 3 March.

Katelaris, A. L., J. Wells, P. Clark, S. Norton, R. Rockett, A. Arnott, V. Sintchenko, S. Corbett and S. K. Bag (2021). “Epidemiologic evidence for airborne transmission of SARS-CoV-2 during church singing, Australia, 2020.” Emerging Infectious Diseases 27(6): 1677.

Korea Centers for Disease, C. and Prevention (2015). “Middle East Respiratory Syndrome Coronavirus Outbreak in the Republic of Korea, 2015.” Osong Public Health and Research Perspectives 6(4): 269–278.

Kucharski, A. and C. L. Althaus (2015). “The role of superspreading in Middle East respiratory syndrome coronavirus (MERS-CoV) transmission.” Eurosurveillance 20(25): 21167.

Lakdawala, S. S. and V. D. Menachery (2021). “Catch me if you can: superspreading of COVID-19.” Trends in Microbiology 29(10): 919–929.

Lam, H. Y., T. S. Lam, C. H. Wong, W. H. Lam, E. L. C. Mei, Y. L. C. Kuen, W. L. T. Wai, B. H. C. Hin, K. H. Wong and S. K. Chuang (2020). “A superspreading event involving a cluster of 14 coronavirus disease 2019 (COVID-19) infections from a family gathering in Hong Kong Special Administrative Region SAR (China).” Western Pacific Surveillance and Response Journal 11(4): 36.

Leo, Y., M. Chen, B. Heng and C. Lee (2003). “Severe acute respiratory syndrome-Singapore, 2003.” MMWR: Morbidity & Mortality Weekly Report 52(18): 405–405.

Leung, K. S.-S., T. T.-L. Ng, A. K.-L. Wu, M. C.-Y. Yau, H.-Y. Lao, M.-P. Choi, K. K.-G. Tam, L. K. Lee, B. K.-C. Wong and A. Y.-M. Ho (2020). “A Territory-wide study of COVID-19 cases and clusters with unknown source in Hong Kong community: A clinical, epidemiological and phylogenomic investigation.” SSRN: preprint.

Lipsitch, M., T. Cohen, B. Cooper, J. M. Robins, S. Ma, L. James, G. Gopalakrishna, S. K. Chew, C. C. Tan and M. H. Samore (2003). “Transmission dynamics and control of severe acute respiratory syndrome.” Science 300(5627): 1966–1970.

Lloyd-Smith, J. O., S. J. Schreiber, P. E. Kopp and W. M. Getz (2005). “Superspreading and the effect of individual variation on disease emergence.” Nature 438(7066): 355–359.

Mahale, P., C. Rothfuss, S. Bly, M. Kelley, S. Bennett, S. L. Huston and S. Robinson (2020). “Multiple COVID-19 outbreaks linked to a wedding reception in rural Maine—August 7–September 14, 2020.” Morbidity and Mortality Weekly Report 69(45): 1686.

Mossong, J., N. Hens, M. Jit, P. Beutels, K. Auranen, R. Mikolajczyk, M. Massari, S. Salmaso, G. S. Tomba and J. Wallinga (2008). “Social contacts and mixing patterns relevant to the spread of infectious diseases.” PLoS Medicine 5(3): e74.

Nishiyama, A., N. Wakasugi, T. Kirikae, T. Quy, L. D. Ha, V. Ban, H. T. Long, N. Keicho, T. Sasazuki and T. Kuratsuji (2008). “Risk factors for SARS infection within hospitals in Hanoi, Vietnam.” Japan Journal of Infectious Diseases 61(5): 388–390.

Sah, P., M. C. Fitzpatrick, C. F. Zimmer, E. Abdollahi, L. Juden-Kelly, S. M. Moghadas, B. H. Singer and A. P. Galvani (2021). “Asymptomatic SARS-CoV-2 infection: A systematic review and meta-analysis.” Proceedings of the National Academy of Sciences 118(34).

Shen, Z., F. Ning, W. Zhou, X. He, C. Lin, D. P. Chin, Z. Zhu and A. Schuchat (2004). “Superspreading SARS events, Beijing, 2003.” Emerging Infectious Diseases 10(2): 256.

Stein, R. A. (2011). “Super-spreaders in infectious diseases.” International Journal of Infectious Diseases 15(8): e510–e513.

Stein, R. A. (2020). “The 2019 coronavirus: Learning curves, lessons, and the weakest link.” International Journal of Clinical Practice 74(4): e13488.

Sugimoto, H. and T. Kohama (2020). “Chest tube with air leaks is a potential “super spreader” of COVID-19.” American Journal of Infection Control 48(8): 969.

Tang, J. W., L. C. Marr, Y. Li and S. J. Dancer (2021). “Covid-19 has redefined airborne transmission.” British Medical Journal 373.

Viceconte, G. and N. Petrosillo (2020). “COVID-19 R0: Magic number or conundrum?” Infectious Disease Reports 12(1).

WHO (2003). “Consensus document on the epidemiology of severe acute respiratory syndrome (SARS).” May 16-17. from

WHO (2017). “WHO MERS-CoV global summary and assessment of risk 2017.” 21 Jul 2017. from

Wilder-Smith, A., M. D. Teleman, B. H. Heng, A. Earnest, A. E. Ling and Y. S. Leo (2005). “Asymptomatic SARS coronavirus infection among healthcare workers, Singapore.” Emerging Infectious Diseases 11(7): 1142.

Woolhouse, M. E., C. Dye, J.-F. Etard, T. Smith, J. Charlwood, G. Garnett, P. Hagan, J. Hii, P. Ndhlovu and R. Quinnell (1997). “Heterogeneities in the transmission of infectious agents: implications for the design of control programs.” Proceedings of the National Academy of Sciences 94(1): 338–342.

World Health Organization (2003). “Consensus document on the epidemiology of severe acute respiratory syndrome (SARS).” May. from https://www.who.int/csr/sars/en/WHOconsensus.pdf.

Xu, X.-K., X. F. Liu, Y. Wu, S. T. Ali, Z. Du, P. Bosetti, E. H. Lau, B. J. Cowling and L. Wang (2020). “Reconstruction of transmission pairs for novel coronavirus disease 2019 (COVID-19) in mainland China: estimation of superspreading events, serial interval, and hazard of infection.” Clinical Infectious Diseases 71(12): 3163–3167.

Yu, X., D. Ran, J. Wang, Y. Qin, R. Liu, X. Shi, Y. Wang, C. Xie, J. Jiang and J. Zhou (2020). “Unclear but present danger: An asymptomatic SARS-CoV-2 carrier.” Genes & Diseases 7(4): 558–566.

## References

Adam, D. C., P. Wu, J. Y. Wong, E. H. Lau, T. K. Tsang, S. Cauchemez, G. M. Leung and B. J. Cowling (2020). “Clustering and superspreading potential of SARS-CoV-2 infec in Hong Kong.” Nature Medicine 26(11): 1714–1719.

Adegboye, O. A. and F. Elfaki (2018). “Network analysis of MERS coronavirus within households, communities, and hospitals to identify most centralized and super-spreading in the Arabian peninsula, 2012 to 2016.” Canadian Journal of Infectious Diseases and Medical Microbiology 2018.

Alanazi, K. H., M. E. Killerby, H. M. Biggs, G. R. Abedi, H. Jokhdar, A. A. Alsharef, M. Mohammed, O. Abdalla, A. Almari and S. Bereagesh (2019). “Scope and extent of healthcare-associated Middle East respiratory syndrome coronavirus transmission during two contemporaneous outbreaks in Riyadh, Saudi Arabia, 2017.” Infection Control & Hospital Epidemiology 40(1): 79–88.

Alenazi, T. H., H. Al Arbash, A. El-Saed, M. M. Alshamrani, H. Baffoe-Bonnie, Y. M. Arabi, S. M. Al Johani, R. e. Hijazi, A. Alothman and H. H. Balkhy (2017). “Identified transmission dynamics of Middle East respiratory syndrome coronavirus infection during an outbreak: implications of an overcrowded emergency department.” Clinical Infectious Diseases 65(4): 675–679.

Assiri, A., G. R. Abedi, A. A. B. Saeed, M. A. Abdalla, M. Al-Masry, A. J. Choudhry, X. Lu, D. D. Erdman, K. Tatti and A. M. Binder (2016). “Multifacility outbreak of middle east respiratory syndrome in Taif, Saudi Arabia.” Emerging Infectious Diseases 22(1): 32.

Atrubin, D., M. Wiese and B. Bohinc (2020). “An outbreak of COVID-19 associated with a recreational hockey game—Florida, June 2020.” Morbidity and Mortality Weekly Report 69(41): 1492.

Barry, M., M. V. Phan, L. Akkielah, F. Al-Majed, A. Alhetheel, A. Somily, S. S. Alsubaie, S. J. McNabb, M. Cotten and A. Zumla (2020). “Nosocomial outbreak of the Middle East Respiratory Syndrome coronavirus: A phylogenetic, epidemiological, clinical and infection control analysis.” Travel Medicine and Infectious Disease 37: 101807.

Bigelow, B. F., O. Tang, G. R. Toci, N. Stracker, F. Sheikh, K. M. J. Slifka, S. A. Novosad, J. A. Jernigan, S. C. Reddy and M. J. Katz (2020). “Transmission of SARS-CoV-2 involving residents receiving dialysis in a nursing home—Maryland, April 2020.” Morbidity and Mortality Weekly Report 69(32): 1089.

Chau, N. V. V., N. T. T. Hong, N. M. Ngoc, T. T. Thanh, P. N. Q. Khanh, L. A. Nguyet, L. N. T. Nhu, N. T. H. Ny, D. N. H. Man and V. T. T. Hang (2021). “Superspreading Event of SARS-CoV-2 Infection at a Bar, Ho Chi Minh City, Vietnam.” Emerging Infectious Diseases 27(1): 310.

Chen, M. I., Y.-S. Leo, B. S. Ang, B.-H. Heng and P. Choo (2006). “The outbreak of SARS at Tan Tock Seng Hospital-Relating epidemiology to control.” Annals of the Academy of Medicine, Singapore 35(5): 317.

Choe, S., H.-S. Kim and S. Lee (2020). “Exploration of superspreading events in 2015 MERS-CoV outbreak in Korea by branching process models.” International Journal of Environmental Research and Public Health 17(17): 6137.

Cooper, B. S., L. Q. Fang, J. P. Zhou, D. Feng, H. Lv, M. T. Wei, S. X. Wang, W. C. Cao and S. J. De Vlas (2009). “Transmission of SARS in three Chinese hospitals.” Tropical Medicine & International Health 14: 71–78.

Danis, K., O. Epaulard, T. Bénet, A. Gaymard, S. Campoy, E. Botelho-Nevers, M. Bouscambert-Duchamp, G. Spaccaferri, F. Ader and A. Mailles (2020a). “Cluster of coronavirus disease 2019 (COVID-19) in the French Alps, February 2020.” Clinical Infectious Diseases 71(15): 825–832.

Danis, K., O. Epaulard, T. Bénet, A. Gaymard, S. Campoy, E. Bothelo-Nevers, M. Bouscambert-Duchamp, G. Spaccaferri, F. Ader and A. Mailles (2020b). “Cluster of coronavirus disease 2019 (Covid-19) in the French Alps, 2020.” Clinical Infectious Diseases(71).

El Bushra, H. E., H. A. Al Arbash, M. Mohammed, O. Abdalla, M. N. Abdallah, Z. K. Al-Mayahi, A. M. Assiri and A. A. BinSaeed (2017). “Outcome of strict implementation of infection prevention control measures during an outbreak of Middle East respiratory syndrome.” American Journal of Infection Control 45(5): 502–507.

Ghinai, I., S. Woods, K. A. Ritger, T. D. McPherson, S. R. Black, L. Sparrow, M. J. Fricchione, J. L. Kerins, M. Pacilli and P. S. Ruestow (2020). “Community transmission of SARS-CoV-2 at two family gatherings—Chicago, Illinois, February–March 2020.” Morbidity and Mortality Weekly Report 69(15): 446.

Goh, K.-T., J. Cutter, B.-H. Heng, S. Ma, B. K. Koh, C. Kwok, C.-M. Toh and S.-K. Chew (2006). “Epidemiology and control of SARS in Singapore.” Annals-Academy of Medicine Singapore 35(5): 301.

Hamner, L. (2020). “High SARS-CoV-2 attack rate following exposure at a choir practice—Skagit County, Washington, March 2020.” MMWR. Morbidity and mortality weekly report 69.

Hijnen, D., A. V. Marzano, K. Eyerich, C. GeurtsvanKessel, A.M. Giménez-Arnau, P. Joly, C. Vestergaard, M. Sticherling and E. Schmidt (2020). “SARS-CoV-2 transmission from presymptomatic meeting attendee, Germany.” Emerging Infectious Diseases 26(8): 1935.

Ho, A. S., J. J. Sung and M. Chan-Yeung (2003). “An outbreak of severe acute respiratory syndrome among hospital workers in a community hospital in Hong Kong.” Annals of Internal Medicine 139(7): 564–567.

Hu, K., Y. Zhao, M. Wang, Q. Zeng, X. Wang, M. Wang, Z. Zheng, X. Li, Y. Zhang and T. Wang (2021). “Identification of a Super-Spreading Chain of Transmission Associated with COVID-19 at the Early Stage of the Disease Outbreak in Wuhan.” Archives of Clinical and Biomedical Research 5(4): 598–612.

Hunter, J. C., D. Nguyen, B. Aden, Z. Al Bandar, W. Al Dhaheri, K. A. Elkheir, A. Khudair, M. Al Mulla, F. El Saleh and H. Imambaccus (2016). “Transmission of Middle East respiratory syndrome coronavirus infections in healthcare settings, Abu Dhabi.” Emerging Infectious Diseases 22(4): 647.

Ismail, S. A., V. Saliba, J. L. Bernal, M. E. Ramsay and S. N. Ladhani (2021). “SARS-CoV-2 infection and transmission in educational settings: a prospective, cross-sectional analysis of infection clusters and outbreaks in England.” The Lancet Infectious Diseases 21(3): 344–353.

Jang, S., S. H. Han and J.-Y. Rhee (2020). “Cluster of coronavirus disease associated with fitness dance classes, South Korea.” Emerging Infectious Diseases 26(8): 1917.

Jiang, S., L. Huang, X. Chen, J. Wang, W. Wu, S. Yin, W. Chen, J. Zhan, L. Yan and L. Ma (2003). “Ventilation of wards and nosocomial outbreak of severe acute respiratory syndrome among healthcare workers.” Chinese Medical Journal 116(9): 1293–1297.

Kang, Y.-J. (2020). “Characteristics of the COVID-19 outbreak in Korea from the mass infection perspective.” Journal of Preventive Medicine and Public Health 53(3): 168.

Khanh, N. C., P. Q. Thai, H.-L. Quach, N.-A. H. Thi, P. C. Dinh, T. N. Duong, L. T. Q. Mai, N. D. Nghia, T. A. Tu and L. N. Quang (2020). “Transmission of SARS-CoV 2 during long-haul flight.” Emerging Infectious Diseases 26(11): 2617.

Kim, K. M., M. Ki, S.-i. Cho, M. Sung, J. K. Hong, H.-K. Cheong, J.-H. Kim, S.-E. Lee, C. Lee and K.-J. Lee (2015). “Epidemiologic features of the first MERS outbreak in Korea: focus on Pyeongtaek St. Mary’s Hospital.” Epidemiology and health 37.

Kim, S. W., J. W. Park, H.-D. Jung, J.-S. Yang, Y.-S. Park, C. Lee, K. M. Kim, K.-J. Lee, D. Kwon and Y. J. Hur (2017). “Risk factors for transmission of Middle East respiratory syndrome coronavirus infection during the 2015 outbreak in South Korea.” Clinical Infectious Diseases 64(5): 551–557.

Kim, Y. and X. Jiang (2020). “Evolving transmission network dynamics of COVID-19 cluster infections in South Korea: a descriptive study.” medRxiv.

Kristiansen, M. F., B. H. Heimustovu, S. áBorg, T. H. Mohr, H. Gislason, L. F. Møller, D. H. Christiansen, B. áSteig, M. S. Petersen and M. Strøm (2021). “Epidemiology and clinical course of first wave coronavirus disease cases, Faroe Islands.” Emerging Infectious Diseases 27(3): 749.

Lee, N., D. Hui, A. Wu, P. Chan, P. Cameron, G. M. Joynt, A. Ahuja, M. Y. Yung, C. Leung and K. To (2003). “A major outbreak of severe acute respiratory syndrome in Hong Kong.” New England Journal of Medicine 348(20): 1986–1994.

Li, Y.-K., S. Peng, L.-Q. Li, Q. Wang, W. Ping, N. Zhang and X.-N. Fu (2020). “Clinical and transmission characteristics of Covid-19 -A retrospective study of 25 cases from a single thoracic surgery department.” Current Medical Science 40(2): 295–300.

Liang, W., Z. Zhu, J. Guo, Z. Liu, X. He, W. Zhou, D. P. Chin, A. Schuchat and B. J. S. E. Group (2004). “Severe acute respiratory syndrome, Beijing, 2003.” Emerging Infectious Diseases 10(1): 25.

Lin, J., K. Yan, J. Zhang, T. Cai and J. Zheng (2020). “A super-spreader of COVID-19 in Ningbo city in China.” Journal of Infection and Public Health 13(7): 935–937.

Liu, Y., R. M. Eggo and A. J. Kucharski (2020). “Secondary attack rate and superspreading events for SARS-CoV-2.” The Lancet 395(10227): e47.

Lu, J., J. Gu, K. Li, C. Xu, W. Su, Z. Lai, D. Zhou, C. Yu, B. Xu and Z. Yang (2020). “COVID-19 outbreak associated with air conditioning in restaurant, Guangzhou, China, 2020.” Emerging Infectious Diseases 26(7): 1628.

Luo, K., Z. Lei, Z. Hai, S. Xiao, J. Rui, H. Yang, X. Jing, H. Wang, Z. Xie and P. Luo (2020). Transmission of SARS-CoV-2 in public transportation vehicles: a case study in Hunan province, China. Open Forum Infectious Diseases, Oxford University Press US.

Mahale, P., C. Rothfuss, S. Bly, M. Kelley, S. Bennett, S. L. Huston and S. Robinson (2020). “Multiple COVID-19 outbreaks linked to a wedding reception in rural Maine— August 7–September 14, 2020.” Morbidity and Mortality Weekly Report 69(45): 1686.

Martinez-Fierro, M. L., J. Ríos-Jasso, I. Garza-Veloz, L. Reyes-Veyna, R. M. Cerda-Luna, I. Duque-Jara, M. Galvan-Jimenez, L. A. Ramirez-Hernandez, A. Morales-Esquivel and Y. Ortiz-Castro (2021). “The role of close contacts of COVID-19 patients in the SARS-CoV-2 transmission: an emphasis on the percentage of nonevaluated positivity in Mexico.” American Journal of Infection Control 49(1): 15–20.

Mohindra, R., A. Ghai, R. Brar, N. Khandelwal, M. Biswal, V. Suri, K. Goyal, M. P. Singh, A. Bhalla and K. Rana (2021). “Superspreaders: A Lurking Danger in the Community.” Journal of Primary Care & Community Health 12: 2150132720987432.

Müller, L., P. N. Ostermann, A. Walker, T. Wienemann, A. Mertens, O. Adams, M. Andree, S. Hauka, N. Lübke and V. Keitel (2021). “Sensitivity of anti-SARS-CoV-2 serological assays in a high-prevalence setting.” European Journal of Clinical Microbiology & Infectious Diseases 40(5): 1063–1071.

Murphy, N., M. Boland, N. Bambury, M. Fitzgerald, L. Comerford, N. Dever, M. B. O’Sullivan, N. Petty-Saphon, R. Kiernan and M. Jensen (2020). “A large national outbreak of COVID-19 linked to air travel, Ireland, summer 2020.” Eurosurveillance 25(42): 2001624.

Nishiura, H., A. Endo, M. Saitoh, R. Kinoshita, R. Ueno, S. Nakaoka, Y. Miyamatsu, Y. Dong, G. Chowell and K. Mizumoto (2016). “Identifying determinants of heterogeneous transmission dynamics of the Middle East respiratory syndrome (MERS) outbreak in the Republic of Korea, 2015: a retrospective epidemiological analysis.” BMJ Open 6(2): e009936.

Ofner-Agostini, M., D. Gravel, L. C. McDonald, M. Lem, S. Sarwal, A. McGeer, K. Green, M. Vearncombe, V. Roth and S. Paton (2006). “Cluster of cases of severe acute respiratory syndrome among Toronto healthcare workers after implementation of infection control precautions: a case series.” Infection Control & Hospital Epidemiology 27(5): 473–478.

Ofner, M., M. Lem, S. Sarwal, M. Vearncombe and A. Simor (2003). “Cluster of severe acute respiratory syndrome cases among protected health-care workers-Toronto, Canada, April 2003.” Morbidity & Mortality Weekly Report 52(19): 433–433.

Olsen, S. J., H.-L. Chang, T. Y.-Y. Cheung, A. F.-Y. Tang, T. L. Fisk, S. P.-L. Ooi, H.-W. Kuo, D. D.-S. Jiang, K.-T. Chen and J. Lando (2003). “Transmission of the severe acute respiratory syndrome on aircraft.” New England Journal of Medicine 349(25): 2416–2422.

Park, G. E., J.-H. Ko, K. R. Peck, J. Y. Lee, J. Y. Lee, S. Y. Cho, Y. E. Ha, C.-I. Kang, J.-M. Kang and Y.-J. Kim (2016). “Control of an outbreak of Middle East respiratory syndrome in a tertiary hospital in Korea.” Annals of internal medicine 165(2): 87–93.

Park, J. Y. (2020). “Spatial visualization of cluster-specific Covid-19 transmission network in South Korea during the early epidemic phase.” medRxiv.

Park, S. Y., Y.-M. Kim, S. Yi, S. Lee, B.-J. Na, C. B. Kim, J.-i. Kim, H. S. Kim, Y. B. Kim and Y. Park (2020). “Coronavirus disease outbreak in call center, South Korea.” Emerging Infectious Diseases 26(8): 1666.

Phiriyasart, F., S. Chantutanon, F. Salaeh, A. Roka, T. Thepparat, S. Kaesaman, A. Useng, I. Abu, R. Weruma and S. Arrong (2020). “Outbreak investigation of coronavirus disease (COVID-19) among islamic missionaries in Southern Thailand, April 2020.” OSIR Journal 13(2).

Phucharoen, C., N. Sangkaew and K. Stosic (2020). “The characteristics of COVID-19 transmission from case to high-risk contact, a statistical analysis from contact tracing data.” EClinicalMedicine 27: 100543.

Poutanen, S. M., D. E. Low, B. Henry, S. Finkelstein, D. Rose, K. Green, R. Tellier, R. Draker, D. Adachi and M. Ayers (2003). “Identification of severe acute respiratory syndrome in Canada.” New England Journal of Medicine 348(20): 1995–2005.

Prevention, C. f. D. C. (2003). “Severe acute respiratory syndrome--Taiwan, 2003.” Morbidity and mortality weekly report 52(20): 461–466.

Saad, M., A. S. Omrani, K. Baig, A. Bahloul, F. Elzein, M. A. Matin, M. A. Selim, M. Al Mutairi, D. Al Nakhli and A. Y. Al Aidaroos (2014). “Clinical aspects and outcomes of 70 patients with Middle East respiratory syndrome coronavirus infection: a single-center experience in Saudi Arabia.” International Journal of Infectious Diseases 29: 301–306.

Safer, M., H. Letaief, A. Hechaichi, C. Harizi, S. Dhaouadi, L. Bouabid, S. Darouiche, D. Gharbi, N. Elmili and H. B. Salah (2021). “Identification of transmission chains and clusters associated with COVID-19 in Tunisia.” BMC Infectious Diseases 21(1): 1–8.

Schwierzeck, V., J. C. König, J. Kühn, A. Mellmann, C.L. Correa-Martínez, H. Omran, M. Konrad, T. Kaiser and S. Kampmeier (2021). “First Reported Nosocomial Outbreak of Severe Acute Respiratory Syndrome Coronavirus 2 in a Pediatric Dialysis Unit.” Clinical Infectious Diseases 72(2): 265–270.

Shen, Y., C. Li, H. Dong, Z. Wang, L. Martinez, Z. Sun, A. Handel, Z. Chen, E. Chen and M. H. Ebell (2020). “Community outbreak investigation of SARS-CoV-2 transmission among bus riders in eastern China.” JAMA Internal Medicine 180(12): 1665–1671.

Shim, E., A. Tariq, W. Choi, Y. Lee and G. Chowell (2020). “Transmission potential and severity of COVID-19 in South Korea.” International Journal of Infectious Diseases 93: 339–344.

Simmerman, J. M., D. Chu and H. Chang (2003). “Implications of unrecognized severe acute respiratory syndrome.” The Nurse Practitioner 28(11): 21–22.

Szablewski, C. M., K. T. Chang, M. M. Brown, V. T. Chu, A. R. Yousaf, N. Anyalechi, P. A. Aryee, H. L. Kirking, M. Lumsden and E. Mayweather (2020). “SARS-CoV-2 transmission and infection among attendees of an overnight camp—Georgia, June 2020.” Morbidity and Mortality Weekly Report 69(31): 1023.

Thompson, E. R., F. S. Williams, P. A. Giacin, S. Drummond, E. Brown, M. Nalick, Q. Wang, J. R. McDonald and A. L. Carlson (2021). “Universal masking to control healthcare-associated transmission of severe acute respiratory coronavirus virus 2 (SARS-CoV-2).” Infection Control & Hospital Epidemiology: 1–7.

Thway, A. M., H. Tayza, T. T. Win, Y. M. Tun, M. M. Aung, Y. N. Win and K. M. Tun (2020). “Epidemiological characteristics of SARS-COV-2 in Myanmar.” medRxiv.

Tian, S., M. Wu, Z. Chang, Y. Wang, G. Zhou, W. Zhang, J. Xing, H. Tian, X. Zhang and X. Zou (2021). “Epidemiological investigation and intergenerational clinical characteristics of 24 coronavirus disease patients associated with a supermarket cluster: a retrospective study.” BMC Public Health 21(1): 1–9.

Tsang, K. W., P. L. Ho, G. C. Ooi, W. K. Yee, T. Wang, M. Chan-Yeung, W. K. Lam, W. H. Seto, L. Y. Yam and T. M. Cheung (2003a). “A cluster of cases of severe acute respiratory syndrome in Hong Kong.” New England Journal of Medicine 348(20): 1977–1985.

Tsang, T., T. Lai-Yin, L. Pak-Yin and M. Lee (2003b). “Update: outbreak of severe acute respiratory syndrome-worldwide, 2003.” Morbidity & Mortality Weekly Report 52(12): 241–241.

Valent, F., T. Gallo, E. Mazzolini, C. Pipan, A. Sartor, M. Merelli, G. Bontempo, S. Marzinotto, F. Curcio and C. Tascini (2020). “A cluster of COVID-19 cases in a small Italian town: a successful example of contact tracing and swab collection.” Clinical Microbiology and Infection 26(8): 1112–1114.

Varia, M., S. Wilson, S. Sarwal, A. McGeer, E. Gournis and E. Galanis (2003). “Investigation of a nosocomial outbreak of severe acute respiratory syndrome (SARS) in Toronto, Canada.” Canadian Medical Association Journal 169(4): 285–292.

Viner, R. M., O. T. Mytton, C. Bonell, G. Melendez-Torres, J. L. Ward, L. Hudson, C. Waddington, J. Thomas, S. Russell and F. van der Klis (2020). “Susceptibility to and transmission of COVID-19 amongst children and adolescents compared with adults: a systematic review and meta-analysis.” medRxiv: 27.

Wei, C., Y. Yuan and Z. Cheng (2020). “A super-spreader of SARS-CoV-2 in incubation period among health-care workers.” Respiratory Research 21(1): 1–3.

Weissberg, D., J. Böni, S. K. Rampini, V. Kufner, M. Zaheri, P. W. Schreiber, I. A. Abela, M. Huber, H. Sax and A. Wolfensberger (2020). “Does respiratory co-infection facilitate dispersal of SARS-CoV-2? investigation of a super-spreading event in an open-space office.” Antimicrobial Resistance & Infection Control 9(1): 1–8.

WHO (2017). “WHO MERS-CoV global summary and assessment of risk 2017.” 21 Jul 2017. from

Wong, R. S. and D. S. Hui (2004). “Index patient and SARS outbreak in Hong Kong.” Emerging Infectious Diseases 10(2): 339.

Xu, X.-K., X.-F. Liu, L. Wang, S. T. Ali, Z. Du, P. Bosetti, B. J. Cowling and Y. Wu (2020a). “Household transmissions of SARS-CoV-2 in the time of unprecedented travel lockdown in China.” medRxiv.

Xu, X.-K., X. F. Liu, Y. Wu, S. T. Ali, Z. Du, P. Bosetti, E. H. Lau, B. J. Cowling and L. Wang (2020b). “Reconstruction of transmission pairs for novel coronavirus disease 2019 (COVID-19) in mainland China: estimation of superspreading events, serial interval, and hazard of infection.” Clinical Infectious Diseases 71(12): 3163–3167.

Yin, G. and H. Jin (2020). “Comparison of transmissibility of coronavirus between symptomatic and asymptomatic patients: reanalysis of the Ningbo COVID-19 data.” JMIR Public Health and Surveillance 6(2): e19464.

Yusef, D., W. Hayajneh, S. Awad, S. Momany, B. Khassawneh, S. Samrah, B. Obeidat, L. Raffee, I. Al-Faouri and A. B. Issa (2020). “Large outbreak of coronavirus disease among wedding attendees, Jordan.” Emerging Infectious Diseases 26(9): 2165.

Zeng, G., S.-Y. Xie, Q. Li and J.-M. Ou (2009). “Infectivity of severe acute respiratory syndrome during its incubation period.” Biomedical and Environmental Sciences 22(6): 502–510.

Zhang, J., P. Zhou, D. Han, W. Wang, C. Cui, R. Zhou, K. Xu, L. Liu, X. Wang and X. Bai (2020). “Investigation on a cluster epidemic of COVID-19 in a supermarket in Liaocheng, Shandong province.” Zhonghua liuxingbingxue zazhi 41: E055–E055.

Zhong, N., B. Zheng, Y. Li, L. Poon, Z. Xie, K. Chan, P. Li, S. Tan, Q. Chang and J. Xie (2003). “Epidemiology and cause of severe acute respiratory syndrome (SARS) in Guangdong, People’s Republic of China, in February, 2003.” The Lancet 362(9393): 1353–1358.

